# Assessment of upper limb movement disorders using wearable sensors during functional tasks: a systematic review

**DOI:** 10.1101/2022.10.13.22281023

**Authors:** Inti Vanmechelen, Helga Haberfehlner, Joni De Vleeschhauwer, Ellen Van Wonterghem, Hilde Feys, Kaat Desloovere, Jean-Marie Aerts, Elegast Monbaliu

## Abstract

**Background:** Studies aiming to objectively quantify upper limb movement disorders during functional tasks using wearable sensors have recently increased, but there is a wide variety in described measurement and analyzing methods, hampering standardization of methods in research and clinics. Therefore, the primary objective of this review was to provide an overview of sensor set-up and type, included tasks, sensor features and methods used to quantify movement disorders during upper limb tasks in multiple pathological populations. The secondary objective was to select the most sensitive sensor features for symptom detection and quantification and discuss application of the proposed methods in clinical practice.

**Methods:** A literature search using Scopus, Web of Science, and PubMed was performed. Articles needed to meet following criteria: (1) participants were adults/children with a neurological disease, (2) (at least) one sensor was placed on the upper limb for evaluation of movement disorders during functional tasks, (3) comparisons between: groups with/without movement disorders, sensor features before/after intervention, or sensor features with a clinical scale for assessment of the movement disorder. (4) Outcome measures included sensor features from acceleration/angular velocity signals.

**Results:** A total of 101 articles were included, of which 56 researched Parkinson’s Disease. Wrist(s), hand and index finger were the most popular sensor locations. The most frequent tasks for assessment were: finger tapping, wrist pro/supination, keeping the arms extended in front of the body and finger-to-nose. The most frequently calculated sensor features were mean, standard deviation, root-mean-square, ranges, skewness, kurtosis and entropy of acceleration and/or angular velocity, in combination with dominant frequencies and power of acceleration signals. Examples of clinical applications were automatization of a clinical scale or discrimination between a patient/control group or different patient groups.

**Conclusion:** Current overview can support clinicians and researchers to select the most sensitive pathology-dependent sensor features and measurement methodologies for detection and quantification of upper limb movement disorders and for the objective evaluations of treatment effects. The insights from Parkinson’s Disease studies can accelerate the development of wearable sensors protocols in the remaining pathologies, provided that there is sufficient attention for the standardisation of protocols, tasks, feasibility and data analysis methods.

## 1 Introduction

The execution of functional tasks requires fine-tuned coordination of multiple upper limb joints, which is often disturbed in individuals with movement disorders [1-3]. Movement disorders can be defined as “a neurological syndrome in which there is either an excess of movement or a paucity of voluntary and automatic movements” and are the consequence of lesions in the basal ganglia, cerebellum or thalamus brain regions. They are present in a variety of neurological diseases and can occur in every phase of the life cycle [4]. Prevalence of movement disorders increases with age, up to 28% in a general population over 50 years old and 50% for individuals over 80 years old [5]. In several neurologic diseases, movement disorders belong to the main symptom of the disease. In childhood, neurologic movement disorders are most often associated with a diagnosis of dyskinetic cerebral palsy (CP) or with primary dystonias (i.e., inherited or idiopathic dystonias) with a prevalence of 25-50/100 000 and 15-30/100 000, respectively [6-8]. In individuals over the age of 50 years, the prevalence of primary dystonia increases to 732/100 000 [9]. In the elderly, the most prevalent condition causing movement disorders is Parkinson’s disease (PD), reporting a prevalence of 1-2 per 1000 adults [10].

Movement disorders lead to slower movement execution, increased movement variability and a decrease in functionality [11-15]. Both in early-onset and late-onset movement disorders, accurate evaluation is indispensable for the follow-up of the disease course – especially in progressive movement disorders – and to evaluate and optimize the effect of treatment strategies. Currently, the effect of an intervention program on upper limb function or the presence and/or severity of movement disorders is mostly evaluated using clinical assessment scales such as functional scales and movement disorder severity scales [16-18]. The Unified Parkinson’s Disease Rating Scale (UPDRS), the Movement Disorders Society revised version of this scale (MDS-UPDRS) and the Hoehn and Yahr scales are currently the most often used assessment scales in PD, whereas the Essential Tremor Rating Assessment Scale is used to rate the severity of essential tremor during nine functional tasks [19-22]. To evaluate the severity of ataxia, the Scale for the Assessment and Rating of Ataxia (SARA) is most often applied [23]. In stroke, the Wolf Motor Function Test (WMFT) and Fugl-Meyer Assessment (FMA) are mainly used to evaluate motor function post-stroke [19, 21, 24-26]. The Action Research Arm Test (ARAT), Box & Block test, Nine Hole Peg Test and Jebsen-Taylor Test evaluate hand function in multiple pathologies, amongst other multiple sclerosis (MS) and stroke, whereas the Monkey Box test was recently developed to evaluate bilateral motor function in Huntington’s Disease (HD) [27-29]. For children with CP, the Melbourne Assessment is a validated measure for upper limb activity [30, 31]. Apart from upper limb activity evaluation scales, the severity of movement disorders such as dystonia can be evaluated with the Burke–Fahn–Marsden Dystonia Rating Scale (BFMDRS) or the Dyskinesia Impairment Scale (DIS) in children and adolescents with dyskinetic CP [32, 33].

A common drawback of all abovementioned activity and movement disorder severity assessment scales is that they have to be evaluated by clinicians through the use of standardized guidelines or definitions with respect to task execution or presence/severity of the movement disorder. This clinical judgement induces subjectivity, as not all clinicians may interpret a definition or guideline in exactly the same manner. Moreover, the attribution of scores by a clinician based on video recordings is time-consuming, especially if frequent monitoring is required to evaluate disease progression or the effect of an intervention.

In an effort to reduce the subjective aspect in the evaluation of movement disorders, motion analysis has been widely introduced as an alternative to objectify movement disorders, as well as to evaluate the effect of treatment interventions in PD [34, 35], CP [36-38] and stroke [39-41]. While three-dimensional motion analysis is the gold standard in movement analysis, it requires a specially equipped expensive laboratory whereby patients need to visit the hospital or study center for study participation or assessment of rehabilitation.

With both the time-consuming aspect of clinical scoring and the location-restricted aspect of three-dimensional motion analysis as main drivers, multiple studies have recently attempted to automate clinical scales with the use of wearable sensors or inertial measurement units (IMUs). These devices are attractive because of their ease-of-use and portability, omitting the necessity for a standardized laboratory which is in particular relevant for long-time follow-up or home-based measures for less mobile patients. IMUs measure linear acceleration and angular velocity of the segment they are placed on, whereas accelerometers measure only acceleration and gyroscopes measure only angular velocity. Specific features derived from acceleration and angular velocity measures can be used to characterize (pathological) movement patterns during multiple tasks or daily life activities. The use of wearable sensors for objective assessment has been previously discussed in PD [42], but this overview focused on all symptoms of PD, consequently providing very little information on specific upper limb tasks. Similarly, Tortelli and colleagues discussed the use of portable digital sensors in HD, whereby the focus was mostly on the assessment of activity and gait [43]. In dyskinetic CP, a recent review discussed instrumented measures for the assessment of dyskinetic CP symptoms, but this scope was not limited to IMUs and therefore less detailed on the topic [44]. While these previous reviews provide much needed insights in the domain of each pathology, an overarching view of sensor protocols and features for the assessment of movement disorders during upper limb tasks could enhance standardisation of data collection. Such standardisation facilitates multi-centre studies and international collaborations and comparison between characteristics of movement disorders between diseases. Therefore, the primary objective of this review was to provide an overview of sensor set-up and type, included tasks, sensor features and methods that are used to evaluate movement disorders during upper limb tasks in multiple pathological populations. The secondary objective was to select the most sensitive sensor features for symptom detection and quantification and describe the application of the proposed methods in clinical practice.

## 2 Methods

### Search strategy

The full literature search was conducted following the Preferred Reporting Items for Systematic Reviews and Meta-Analyses (PRISMA) guidelines [45]. A literature search using three different databases was performed: Scopus, Web of Science, and PubMed until July 2022. Following terms were used in “all fields”:

#1: sensor OR inertial measurement unit; #2: arm OR upper limb; #3: movement disorder

Subsequently, all three databases were searched for #1 AND #2 AND #3.

### Article screening

Articles (n = 990) retrieved from the literature search were extracted. An overview of the articles retained at each stage of the screening process can be found in the PRISMA flow diagram presented in Figure 1 [45]. Any duplicated articles, retrieved by more than one database, were removed by de-duplication based on congruity in authors, title, and year of publication.

**Figure 1:**
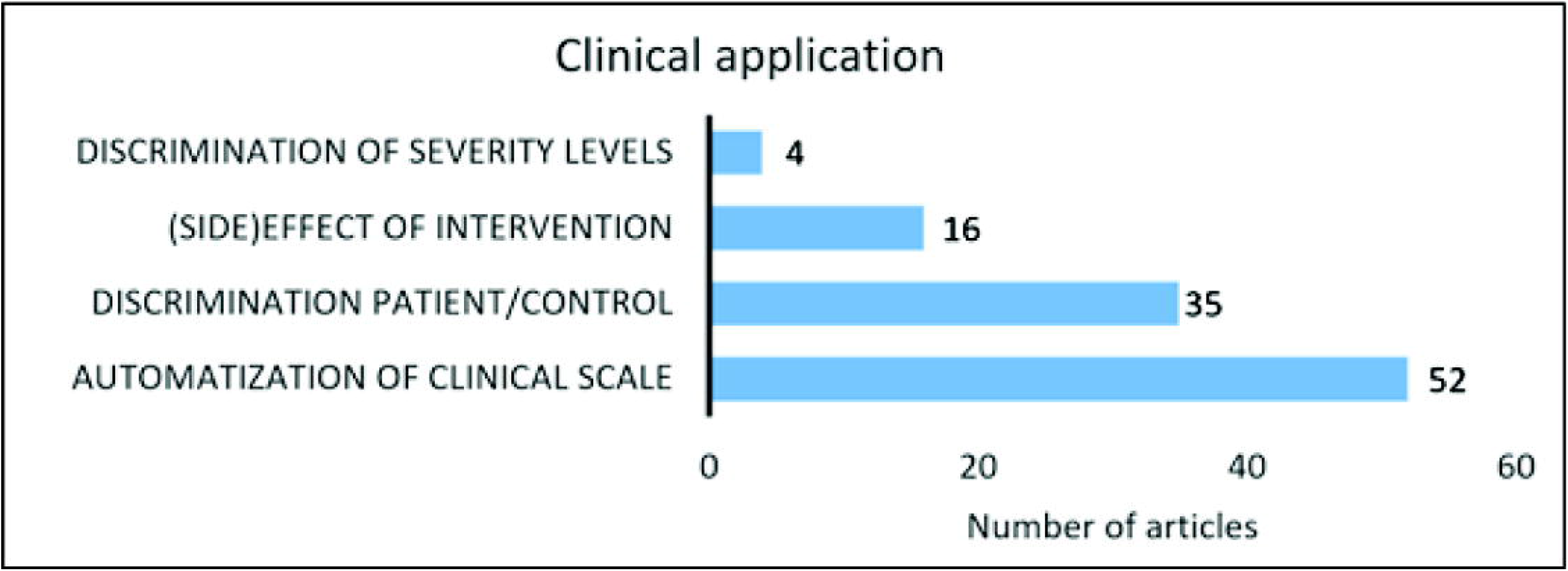
Flowchart of article selection

Unique articles (n = 903) were screened by I.V. for inclusion according to the criteria below in two consecutive stages: (1) title-abstract; and (2) full-text screening.

Articles were screened for inclusion along a set of pre-defined eligibility criteria for (1) the title-abstract and (2) the full-text screening stages. These criteria were designed in line with the PICO/PECO framework [46], which clarifies the review objectives and inclusion criteria across four domains: (P) it was required that the participants were adults or children with a neurological disease subsequently leading to a movement disorder in (but not limited to) the upper limb. (I/E) a minimum of one wearable sensor was placed on the upper limb for the evaluation of movement disorders during the execution of an upper limb task. (C) multiple comparisons were possible: i) a group with movement disorders compared with a healthy group, ii) comparison of sensor features before and after an intervention or iii) comparison of sensor features with scores of a clinical scale. (O) Outcome measures needed to include sensor features derived from acceleration or angular velocity signals. Studies from the same authors who mentioned the exact same features in the same population as a study that was already included were excluded. Additionally, to meet the inclusion criteria, articles were required to be original research containing empirical data. Finally, only articles published after the year 2000 were included.

### Data extraction

Relevant information from each included article was extracted in a custom-made Excel based (Microsoft Office, Microsoft, Redmond, WA, USA) data extraction form. Information regarding goal population, sensor type, number of sensors, location of sensor(s), included tasks, sensor features and statistical method was obtained to address objective 1. To address objective 2, the sensitivity and/or responsiveness of the sensor features were extracted for the articles that provided the contribution of individual sensor features. Finally, the clinical application of the proposed method was extracted.

## 3 Results

### General information

From the 166 full-text articles screened for eligibility, 62 were finally included. Additionally, 39 articles were included from citations of screened articles. The full-text articles that were screened but excluded and the reasons for exclusion can be found in Supplementary Material S1.

Of the included studies, 56 included adults with PD [11, 47-101], of which 46 assessed one or multiple symptoms of PD and 10 studies specifically focused on Parkinsonian tremor [91-100]. Twelve studies included patients with essential tremor [102-113] and 11 included adults post-stroke [15, 28, 114-122], whereas six included adults with MS [123-128]. One study included adults with HD and eight studies included children or adults with ataxia [29, 129-136]. Five studies included children with CP while two studies included children with dystonia and spasticity, respectively [12, 13, 137-141] (Figure 2).

**Figure 2:**
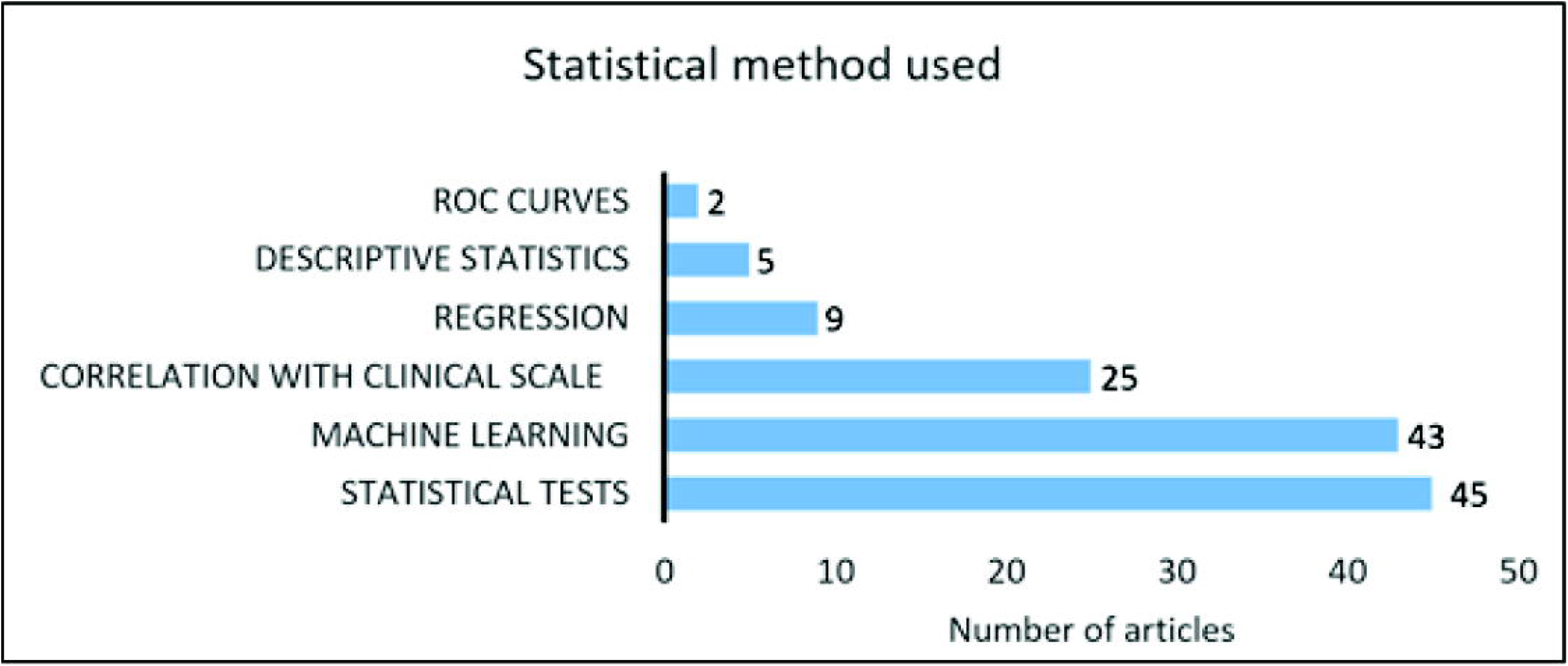
Number of studies included per goal population

### Type, number and location of sensors

Table 1 provides an overview of type and number of sensors used, and their respective location for all included studies. From the 101 identified studies, 24 studies used an accelerometer [29, 47-49, 51, 53, 54, 56, 63, 89, 91, 94, 96, 104, 111, 115, 116, 119, 120, 127, 134, 138, 140, 142], 13 studies measured motion with a gyroscope or angular sensor [50, 52, 58-61, 64, 73, 79, 105, 108, 110], four studies collected motion data using an orientation sensor, motion sensor or magnetic motion tracker [13, 55, 80, 114], 58 studies used IMUs including an accelerometer and gyroscope [11, 12, 15, 28, 57, 62, 65, 67-72, 74-78, 81-88, 90, 93, 98-101, 106, 107, 109, 112, 113, 117, 118, 121-123, 125, 126, 128, 129, 131-137, 139, 141, 143, 144] and three studies included IMUs but only used the acceleration signal for further analysis [102, 103, 107], while one study used IMUs but only processed angular velocity signals [97].

**Table 1:** Number, type and location of included sensors per reference. † Either 1 sensor was placed on the dorsal hand, or 1 on each hand. * Krishna et al. used one sensor but placed it subsequently on the L/R wrist, L/R dorsal hand and L/R ankle.

| # of sensor(s) | Location of sensor(s) | Type | Goal population |
| --- | --- | --- | --- |
| 1 | Index finger tip | Accelerometer | PD [51, 91, 94, 97] |
| 1 | Index finger tip | Gyroscope | PD [59, 61] |
| 1 | Index finger | Motion sensor | PD [69] |
| 1 | Index finger | Inertial measurement unit | PD [99], tremor [113] |
| 1 | Dorsal hand | Angular sensor | PD [50, 79] |
| 1 | Dorsal hand | Orientation sensor | Stroke [114] |
| 1 | Dorsal hand | Accelerometer | Tremor [104] |
| 1 | Wrist | Accelerometer | PD [63, 89, 96], dystonia [140], ataxia [134], MS [127] |
| 1 | Forearm | Inertial measurement unit | PD [72], stroke [117] |
| 1/2 [66]† | Dorsal hand | Inertial measurement unit | PD [66, 68, 75, 84], ataxia [131], tremor [106, 109] |
| 1/2 [78, 81, 82, 133, 137]† | Wrist(s) | Inertial measurement unit | PD [78, 81, 82, 100], stroke [15, 118, 122], CP [137], MS [123, 124, 126], ataxia [133], tremor [112] |
| 1/2 [60]† | Wrist(s) | Gyroscope | PD [52, 60], tremor [108] |
| 2 | Hand and wrist | Inertial measurement unit & accelerometer | PD [86], ataxia [144] |
| 2 | Dorsal hand and index finger | Inertial measurement unit | PD [135] |
| 2 | Wrist and index finger | Accelerometer | PD [67, 142], tremor [111] |
| 2 | Thumb and index finger | Accelerometer | PD [54] |
| 2 | Thumb and index finger | Gyroscope | PD [64, 73] |
| 2 | Thumb and index finger | Inertial measurement unit | PD [58, 70, 71, 85, 88, 101] |
| 3 | Index finger, forearm, upper arm | Inertial measurement unit | Ataxia [132, 135] |
| 3 | Index finger, hand, forearm | Gyroscope | Tremor [105, 110] |
| 3 | Hand, forearm, upper arm | Inertial measurement unit | PD [74, 92], stroke [116], tremor [102] |
| 3 | Wrists and sternum | Accelerometer | HD [29] |
| 3 | Wrist, sternum and sacrum | Inertial measurement unit | Stroke [121] |
| 3 | Upper arms and trunk | Inertial measurement unit | CP [12] |
| 4 | Thumb, index finger, wrist, upper arm | Inertial measurement unit | Stroke [119] |
| 4 | Fingers and wrist | Inertial measurement unit | PD [83] |
| 4 | Hands and forearms | Inertial measurement unit | PD [76], tremor [103] |
| 4 | Wrists, trunk, head | Inertial measurement unit | PD [90] |
| 4 | Wrists and ankles | Inertial measurement unit | PD [77, 87], dyskinetic CP [139] |
| 4 | Wrist, sternum, thigh, foot | Inertial measurement unit | PD [57] |
| 4 | Hand, forearm, upper arm, shoulder | Inertial measurement unit | Spasticity [141] |
| 5 | Thumb, index finger, hand, wrist, upper arm | Magneto-inertial sensors | PD [80] |
| 5 | Hand, wrist, upper arm, shoulder, sternum | Inertial measurement unit | MS [125, 128] |
| 5 | Hand, forearm, upper arm, head, back | Orientation sensors | Dyskinetic CP [13] |
| 6 | Thumb, index finger, hand, forearm, upper arm, sternum | Accelerometer | Stroke [115, 120] |
| 6 | Hands, forearms, upper arms | Inertial measurement unit | PD [98], tremor [107] |
| 6* | Dorsal hands, wrists, ankles | Inertial measurement unit | Ataxia [129] |
| 6 | Hand, scapula, thorax, sacrum, posterior of the head, lateral shank | Magnetic motion tracker | PD [55] |
| 6 | Wrists, waist, chest, ankles | Inertial measurement unit | PD [62] |
| 7 | Hands, wrists, upper arms, sternum | Inertial measurement unit | Stroke [28] |
| 7 | Wrists, upper arms, trunk, upper legs | Accelerometer | PD [47, 48] |
| 8 | Hands, wrists, upper arms, shoulder | Inertial measurement unit | Stroke [65] |
| 8 | Forearms, upper arms, sternum, thighs, right shin | Accelerometer | PD [49] |
| 8 | Forearms, upper arms, shins, upper legs | Accelerometer | PD [53, 56] |
| 8 | index finger, dorsal hand, wrist, dorsum foot, sternum, upper-back, ankles | Inertial measurement unit | Ataxia [136] |
| 11 | Dorsal hand and fingers | Inertial measurement unit | PD [11] |
| 17 | Hands, wrists, upper arms, clavicle's, sternum, head, pelvis, upper & lower legs, feet | Inertial measurement unit | PD [93] |
| Unknown | Upper limb (no specification) | Accelerometer | CP [138] |

The number of sensors ranged from one to 17. Thirty-seven studies used only one sensor, either on the finger, hand, wrist or forearm [15, 50-52, 59, 61, 63, 68, 69, 72, 75, 79, 84, 89, 91, 94, 96, 97, 99, 100, 104, 106, 108, 109, 112-114, 117, 118, 122-124, 126, 127, 131, 134, 140], while seven studies used two sensors bilaterally placed on the hand or wrist [60, 66, 78, 81, 82, 133, 137]. Nine studies used two sensors of which the majority placed one on the thumb and one the index finger [54, 58, 64, 70, 71, 73, 85, 88, 101], while Martinez-Manzara et al. used one sensor on the hand and one on the finger [67], Samotus et al. and Rahimi et al. put one on the wrist and one on the index finger [95, 111] and Shawen et al. put one on the hand and one on the wrist [86]. In the studies where three sensors were used, the most frequent locations for PD and tremor were hand, forearm and upper arm [74, 92, 102, 116], index finger, hand, forearm [105, 110] or index finger, forearm and upper arm [132, 135]. In children with CP, Newman et al. attached one sensor on the sternum and two on both upper arms [12] and in HD, Bennasar et al. placed one sensor on the sternum and two on the wrists [29]. In stroke, Van Meulen et al. fixed one sensor on the wrist, sternum and sacrum [121]. When four sensor were used, sensor placements were: three fingers and the wrist in PD [83], thumb, index finger, wrist and upper arm in stroke [119], hand, forearm, upper arm and shoulder in spasticity [141], hands and forearms [90, 103] and wrists, trunk and head in PD [76]. Four more studies also included lower limb sensors, where three placed sensors on both wrists and ankles and Zwartjes et al. included wrist, foot, thigh and sternum [57, 77, 87, 139]. Four studies used five sensors on the upper limbs, with sensor placement on hand, wrist, upper arm, shoulder and sternum in MS [125, 128], thumb, index finger, hand, forearm, upper arm and sternum for Di Biase et al. and hand forearm, upper arm, head and back for Sanger et al. (note that both studies are based on magnetic or orientation sensors) [13, 80]. Seven studies used six sensors, where the sensor placement was thumb, index finger, hand, forearm, upper arm and sternum [115, 120]. Two studies placed the sensors on hands, forearms and upper arms [98, 107], while Krishna et al. used one sensor but subsequently fixed it on both hands, wrists and ankles, thus including six sensor signals in the analysis [129]. Cheralu et al. placed sensors on the hand, scapula, thorax, sacrum, head and shank, while Tsipouras et al. attached sensors bilaterally on ankles and wrists and one the waist and chest [55, 62]. Repnik et al. used seven sensors on the hands, wrists, upper arms and sternum and Hof et al. and Keijsers et al. placed the sensors on the wrists, upper arms, trunk and upper legs [28, 47, 48]. Five studies used eight sensors. Delrobaei et al. placed the sensors on the hands, wrists, upper arms, and shoulders and Bonato et al. on the forearms, upper arms, thighs, right shin and sternum [49, 65]. Patel et al. and Cole and colleagues attached the sensors on the forearms, upper arms, shins and upper legs [53, 56], whereas in ataxia, Kashyap and colleagues placed sensors on the index finger, hand, wrist, foot, sternum, back and ankles [136]. Finally, Van den Noort et al. used 11 sensors, all located on the hand and fingers [11] and Delrobaei and colleagues used 17 sensors placed on the hands, wrists, upper arms, clavicle’s, sternum, head, pelvis, upper and lower legs and feet [93]. In one study, the number of sensors was not specified [138].

### Upper limb tasks

The upper limb tasks occurring in more than one study are presented in Figure 3. Wrist pro/supination was included in 25 studies [11, 50, 52, 53, 58, 60, 65, 69, 72-74, 76, 78-83, 86, 88, 101, 106, 129, 132, 144] whereas finger tapping was included in 24 studies [11, 49, 51, 54, 58, 59, 61, 64, 67, 69, 71-73, 79, 80, 83, 85, 88, 101, 131, 136, 144-146]. Keeping arms in front of the body was included in 23 studies [52, 66, 91, 93, 95, 97-100, 103-105, 107-113, 125-128], as well as finger-to-nose [13, 53, 76, 86, 91, 97-99, 104, 108, 112, 113, 124-129, 132, 133, 135, 136, 144]. Drinking from a can/cup was included in 13 studies [47, 48, 52, 57, 62, 76, 86, 87, 107, 113, 117, 120, 141], and opening/closing of the hand in seven studies [11, 58, 69, 71, 83, 88, 101] as well as writing/drawing [52, 76, 77, 86, 102, 106, 113]. Eating was included in six studies [48, 52, 68, 77, 84, 87] as well as pouring water [76, 86, 108, 113, 126, 127] whereas reaching/grasping objects was included in five studies [117, 121, 126, 127, 137]. Teeth brushing was included in four studies [52, 68, 77, 84] as well as putting clothes on/off [47, 48, 77, 87]. In stroke, the Wolf Motor Function test or parts of this clinical scale were included four times [115, 118-120] and four studies measured activities in an unrestricted home environment [63, 89, 134, 139]. Combing hair was included in three studies [52, 77, 87] as well as typing and folding laundry [76, 77, 86] and forwards and sideways reaching [12, 114, 116]. Tasks from the ARAT were included in two studies [28, 123]. Finally, following tasks were included once: the monkey box test [29], the box and block test [90], holding a weight with the wrist [96], wrist extension [75], wrist ab/adduction, flexion/extension, elbow flexion/extension and pro/supination [92], and following a bent wire shape with a wand loop [104]. One study included wrist supination/flexion, hand behind back and wrist flexion/pronation [15]. In CP, one study included outwards reaching [13], one included the drinking test, the bean bag test and the nine hole peg test [141] while Strohrmann et al. included turn around cards, pick up small objects, stack dominos, open & close & bottle, use a key, and the nine-hole peg test [137]. Kim et al. included the Jebsen Taylor Hand Function Test, the Quality of Upper Extremity Skills Test and the Box and Blocks Test [138].

**Figure 3:**
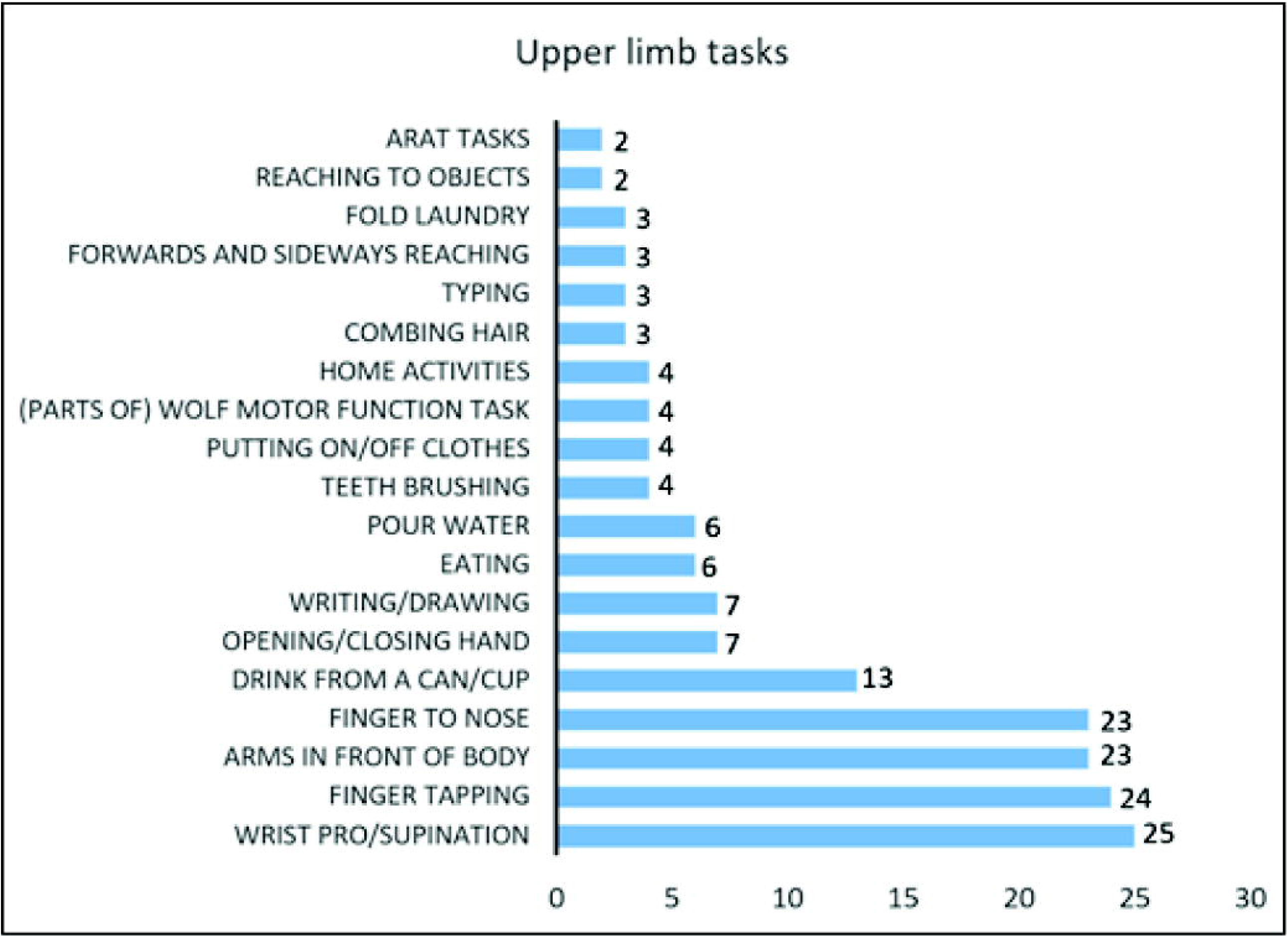
Overview of upper limbs tasks included in more than one study. The sum does not add up to 102 because multiple studies used more than one task.

### Sensor features

Table 2 provides an overview of the calculated sensor features in the time-and frequency domain, as well as a formula or feature description when given in the original study. As an easy and straightforward feature, execution time was often calculated for the upper limb tasks for stroke [15, 28, 117], MS [123], PD [11, 49, 54, 71, 75, 80], tremor [102, 144], CP [137] and ataxia [132, 134]. The frequency of movements was popular in multiple studies in PD, mostly in repetitive tasks such as finger tapping and pro/supination [64, 83].

**Table 2:** Calculated sensor features in time and frequency-domain. STD = standard deviation; max = maximal; RMS = root-mean-square; VAR = variance; IQR = inter-quartile range. PSD = power spectral density

| Time-domain | Formula/Feature description | Goal population |
| --- | --- | --- |
| Execution time | $T_{m_i} = T_{t_i} - T_{0_i}$ [28]; Inter-tap-interval (ITI) [144]; mean, standard deviation, minimum, maximum, range, interquartile range, median, and tenth and ninetieth percentiles of time [133]; duration of sub movements [134] | Stroke [15, 28, 117], PD [11, 49, 54, 71, 75, 80], CP [12, 137], tremor [102, 144], MS [123, 124], ataxia [132, 134] |
| Movement Frequency | Number of rotations/movements [64] | PD [64, 83] |
| Mean acceleration & angular velocity | $m_i(k) = \frac{1}{2sf+1} \sum_{n=w(k)-sf}^{w(k)+sf} x_i(n)$ [62]; Mean absolute value ; Absolute & harmonic mean [139] | PD [62, 78, 86, 90], stroke [118], dyskinetic CP [139], ataxia [129] |
| STD acceleration & angular velocity | $s_i(k) = \sqrt{\frac{1}{2sf+1} \sum_{n=w(k)-sf}^{w(k)+sf} (x_i^f(n) - \bar{x}_i^f)^2}$ [62]; $\sigma(W) = \sqrt{\frac{1}{N} \sum_{i=1}^N (x_i - \mu(W))^2}$, $\mu(W) = mean$ [87] | PD [62, 78, 87] |
| MAX acceleration & angular velocity | Absolute max [75] | PD [75, 89], dyskinetic CP [139] |
| Timing of MAX acceleration & angular velocity | Absolute max [75] | PD [75] |
| RMS acceleration & angular velocity |  | PD [11, 86], dyskinetic CP [139], ataxia [129] |
| VAR acceleration & angular velocity |  | Stroke [118]; PD [86, 89], ataxia [129] |
| Mean acceleration | Mean acceleration in 2min epoch [63] | Stroke [115, 119, 120], PD [57, 63, 66, 96], HD [29], CP [137], tremor [111], ataxia [134] |
| STD acceleration |  | PD [54, 66, 89] HD [29], CP [137], tremor [104], ataxia [134] |
| MAX acceleration | Max acceleration in 2min epoch [63]; Moments of jerk magnitude [89] | PD [63, 89], CP [138], ataxia [133] |
| Mean angular velocity | Hand mobility [52] | PD [52, 64, 71, 74, 81, 83, 85], ataxia [132, 133] |
| STD angular velocity |  | PD [74], ataxia [133] |
| Median angular velocity |  | PD [82] |
| Median acceleration |  | PD [89] |
| MAX linear velocity | Average of maximum velocities [51] | PD [51, 54], stroke [115] |
| RMS acceleration |  | PD [53, 66, 89]; stroke [115, 116] |
| RMS angular velocity | Speed [69]; Mean Intensity (MI) = RMS angular velocity [108] | PD [50, 58-60, 69, 79, 88, 98, 99, 101], tremor [105, 108, 110] |
| Mean angular displacement |  | MS [128] |
| RMS angular displacement/movement amplitude | Amplitude [69] | PD [58-60, 69, 79, 83, 92, 93, 101], tremor [107, 110] |
| Range angular displacement | Hand activity [52] | PD [11, 52, 71, 82, 83, 90] |
| IQR angular displacement |  | PD [82] |
| Range acceleration |  | PD [53, 54, 89], stroke [122] |
| Range acceleration and angular velocity |  | PD [75, 76, 86] |
| Mean amplitude | Average amplitude [51] | PD [51, 64] |
| Range angular velocity | $RAV = \frac{1}{3} \sum_{frontal, vertical, lateral} range(\omega)$ | PD [75, 80], CP [12, 137], ataxia [133] |
| IQR acceleration & angular velocity | Angular velocity [82]; angular acceleration [89] | PD [82, 89] |
| Range of jerk & angular acceleration | $(Acc/Gyr)_{ran} = \max(\dot{x}(t)) - \min(\dot{x}(t)) \cdot t \in \{1, W_n\}$ [72] | PD [72] |
| Peak-to-peak angular velocity | Difference between the mean of the highest and lowest 10 samples in $W$ [87] | PD [73, 80, 87] |
| Magnitude angular velocity |  | PD [77] |
| Max, STD, RMS & min/max peak height |  | HD |
| Rotational jerk index | $rot_i = \log \sqrt{\frac{(T_{t_i} - T_{0_i})^2}{2\theta^p} \int_{T_{0_i}}^{T_{t_i}} \left\ \frac{d^2 \omega(t)}{dt^2} \right\ ^2 dt}$ [28] | Stroke [28] |
| Segment velocity | Square root of sum of squares of jerk signals in three directions [48] | PD [48] |
| Kurtosis | $S(W) = \frac{\frac{1}{N} \sum_{i=1}^N (x_i - \mu(W))^4}{\sigma(W)^4}$ [87] | PD [66, 76, 86, 87, 118], ataxia [134] |
| Skewness | $S(W) = \frac{\frac{1}{N} \sum_{i=1}^N (x_i - \mu(W))^3}{\sigma(W)^3}$ [87] | PD [66, 76, 78, 86, 87, 118] |
| Sample entropy | $H_i(k) = -\frac{1}{2sf+1} \sum_{n=w(k)-sf}^{n=w(k)+sf} p(x_i^f(n)) \log p(x_i^f(n))$ [62] | PD [53, 55, 62, 66, 70, 76, 86], HD [29], stroke [120], ataxia [136] |
| Approximate entropy | $H(W) = -\sum p(B_i) * \log p(B_i)$ [115]; Window length (m) = 2 and % STD (r) = 20% [78] | PD [49, 70, 78, 98, 118], stroke [115, 116] |
| Shannon entropy | Randomness in time domain: $H(W) = -\sum_{i=1}^{200} p(B_i) * \log p(B_i)$ [87] | PD [87], dyskinetic CP [139] |
| Permutation entropy | Assesses the complexity of the time series signal [29] | HD [29] |
| Fuzzy entropy |  | Ataxia [131, 144] |
| Gini index | Movement complexity in the time domain: $G(W) = 1 - \sum_{i=1}^{200} p(B_i)^2$ [87] | PD [87], ataxia [131] |
| Lyapunov exponent | Measures the level of chaos in a signal [29] | HD [29] |
| Recurrence rate (RR); Determinism | RR: Probability that any state will recur again; Determinism: Ratio of recurrence points | HD [29] |
| Average Diagonal line | Average time that signal segments remain the same [29] | HD [29] |

|  |  |  |
| --- | --- | --- |
| RMS of jerk |  | PD [118], stroke [115, 116] |
| Jerk metric | RM jerk normalized by peak velocity [115]; Moments of jerk magnitude [76, 89]; Logarithm of mean jerk amplitude, normalized to mean absolute acceleration movement duration [123]; Normalised Jerk Index: $NJI = \frac{1}{v_{peak}(t_1-t_2)} \int_{t_1}^{t_2} \left\vert \frac{d^2v}{dt^2} \right\vert dt$ [12, 141]; Dimensionless Jerk Index (DLJ) [90]; Mean jerk [15] | Stroke [15, 115], PD [76, 89, 90], Multiple Sclerosis [123], CP [12], dyskinetic CP [13], spasticity [141] |
| Smoothness | Difference between movement accelerometer readings & smoothed readings [122]; Number of movement units [BAI]; number of speed peaks (NSP) [114] | Stroke [122], spasticity [141] |
| Coefficient of variation | Coefficient of variation of amplitude, speed & frequency [64, 71]; STD of a 1-second sliding window of the RMS excursion angle divided by the mean & Coefficient of variation of acceleration & angular velocity [58]; Coefficient of variation of excursion angle [58, 69, 79, 101]; Coefficient of variation of angular velocity [79]; Coefficient of variation of inter-tap - interval [144] | PD [58, 64, 69, 71, 79, 83, 101], tremor [144] |
| Rhythm | STD of intervals a single finger tap movement in 60 sec [51]; Any sequence of regularly recurring events [67]; | PD [51, 67] |
| Variability | RMS error between the reference trial and the warped trial [117] | Stroke [117] |
| Higuchi's fractal dimension (HFD) | Geometrical structure of non-linear time series [12] | CP [12] |
| Correlation between axes | Mean, STD, skewness and kurtosis of signal derivative [86]; Correlation between each two axes of accelerometer [29] | PD [76, 86, 89, 99], HD [29], stroke [115] |
| Peak of normalized cross-correlation from pairs of acceleration time series |  | PD [53] |
| Lag of first peak in autocorrelation acceleration |  | PD [56] |
| Path length | Path-length-ratio (PLR) = distance travelled by hand/straight-line distance [114]; Length of 3D trajectories [121]; Index of curvature [13]; Mean & Standard deviation of Euclidian distance from the mean trajectory [132]; curved line similarity analysis, straight line similarity analysis [135] | Stroke [114, 121], dyskinetic CP [13], spasticity [141], ataxia [132] |
| Similarity of hand trajectories | $D_i = \sqrt{\frac{1}{s_{t_i}-s_{o_i}} \int_{s_{o_i}}^{s_{t_i}} \ p(\bar{s}) - p_r(\bar{s})\ ^2 d\bar{s}}$ , $\hat{v}_i = \frac{1}{\pi} \sqrt{\frac{1}{s_{t_i}-s_{o_i}} \int_{s_{o_i}}^{s_{t_i}} v^2(\hat{s}) d\hat{s}}$ , $X_i = \alpha D_i + \beta \hat{v}_i$ [28] | Stroke [28] |
| Elevation angle | $\theta = \max(\cos^{-1}(\frac{a_{vert(cal)}}{g}))$ [12] | CP [12] |
| Bradykinesia Index (BKI) | $BKI = \sqrt{\frac{STD+VEL}{Time_{var}*Amp_{var}}}$, $var = STD$ of distances between signal peaks for time and amp [65] | PD [65] |
| Movement decrement | Slope of change in amplitude [71]; $\frac{\text{Amplitude in 2nd half time interval}}{\text{Amplitude in 1st half time interval}}$ [69]; Fatigability index: slope of linear equation fitted with peak-to-peak angular velocities [80] | PD [69, 71, 80] |
| Velocity decrement | Compare velocities between 1<sup>st</sup>, 2<sup>nd</sup>, 3<sup>rd</sup> and 4<sup>th</sup> part of the data [82]; $\frac{\text{Speed in 2nd half time interval}}{\text{Speed in 1st half time interval}}$ [69] | PD [69, 82] |
| Amplitude of modulation acceleration |  | PD [49] |
| Normalized mean squared error between a target signal and its forward linear prediction |  | PD [61] |
| **Frequency-domain** |  |  |
| Dominant frequency component | Frequency associated with maximum power [87, 115]; Frequency in 1-4Hz and 4-8Hz bands [47] | PD [47, 53, 84, 87, 91, 94, 97-99, 104, 112, 126, 127, 129], stroke [115] |
| Second dominant frequency | Frequency associated with the second highest peak [87] | PD [87] |
| Dominant frequency of jerk |  | Stroke [115], ataxia [136] |
| Resonant Frequency (FR) | Peaks of FFT waveforms of angles and angular velocity [129] | Ataxia [129, 144] |
| Energy acceleration | Energy in 0.2Hz bin around dominant frequency [15, 115, 119, 120] | Stroke [115, 119, 120], PD [49, 53, 56, 66], HD [29] |
| Energy angular velocity |  | PD [100] |
| Energy acceleration & angular velocity | 2-5Hz: $p_i^l(k) = \sum_{n=w(k)-sf}^{w(k)+sf} (x_i^l(n))^2$; 5-10Hz: $p_i^h(k) = \sum_{n=w(k)-sf}^{w(k)+sf} (x_i^h(n))$; [62] | Stroke [15], PD [62], ataxia [129] |
| Amplitude and dominant frequency of modulation associated with acceleration |  | PD [49] |
| Average magnitude of 1<sup>st</sup> five STFT components |  | HD [29] |
| Fractal dimension acceleration |  | PD [49] |
| Spectral power | Frequency band 1.5-3Hz & 5-8Hz [118]; Frequency band dyskinesia: 0.3-3Hz [77]; Frequency band 0.5-15Hz & 1-4Hz [87]; Frequency band 0.6-16Hz [140]; Welch PSD of displacement; Power Spectral Density plots [68, 84]; PSD ratio 0.5-4Hz & 4-12Hz [102]; logarithm of the peak in the power spectrum in the tremor frequency band [113] | Stroke [118], PD [68, 77, 84, 87], dystonia [140], tremor [102, 113] |
| Peak power | Gyroscope; 0-4Hz frequency band [73]; Power of dominant frequency [87]; Position and amplitude of dominant peaks in power spectra of acceleration signals [104] | PD [59, 60, 73, 87, 99], tremor [102, 104, 105, 110], ataxia [134], MS [125] |
| Second peak power | Power of second dominant frequency [87] | PD [87] |
| Total power | Power spectrum of angular velocity [59]; Gyroscope; 0-4Hz frequency band [73]; power ratio ACC = (Power in 3-7Hz range)/(Power in 3-7Hz+Power in 7-12Hz) [96] | PD [59, 60, 73, 80, 96] |
| Mean power | 0.2-4Hz frequency band [63] | PD [63] |
| Band power |  | Dyskinetic CP [139] |
| Spectral entropy |  | PD [62, 87], Ataxia [131] |
| Component entropy |  | HD [29] |
| Smoothness (Spectral Arc Length) | $SAL_{\pm} = - \int_0^{\Omega_c} \sqrt{\left(\frac{1}{\Omega_c}\right)^2 + \left(\frac{dV(\Omega)}{d\Omega}\right)^2} d\omega$, where $V(\Omega) \pm \frac{V(\Omega)}{V(0)}$ [12] | CP [12], PD [73, 80] |
| Specific tremor index | Tremor Index (TI) = 100*(rms[TR])/(rms[A]) with TR = tremor & A = norm of angular velocity [124]; Average Tremor amplitude (ATA) = $4 * LT_s \sum_{f=f_1}^{f_2} \sqrt{X(f)}$ with L = # of frequency bins, $T_s$ = sampling period, X = signal's PSD [128]; Mean logarithmic tremor power [103]; Tremor rotational amplitude & tremor amplitude based on identified peaks in power spectrum [106] | MS [124, 128], tremor [103, 106] |
| Tremor Frequency (TF) & Tremor Amplitude (TA) | TF= frequency distribution for highest maxima between 1-15Hz; TA = square root of integral of PSD +/- 1Hz of detected frequency [109] | Tremor [109] |

For the studies where both acceleration and angular velocity signals were collected, both mean and standard deviation (STD) were often calculated [62, 78, 86, 87, 90, 118, 129, 139], as well as root-mean-square (RMS) values [11, 86, 89, 129, 139]. Additionally, mean and RMS or STD of acceleration and angular velocity separately were used in studies were one of the signals was available [29, 50, 52, 54, 57, 63, 64, 66, 71, 74, 82, 83, 85, 89, 96, 104, 105, 108, 110, 111, 115, 119, 120, 132-134, 137], as well as median angular velocity in one study [82]. Maximal linear velocity was additionally often used as key feature, mostly by integration of the acceleration signal [51, 54, 115]. The range of angular displacement or range of motion was only used in studies in PD [58-60, 69, 79, 83], mainly to assess hypokinesia. The range of acceleration and angular velocity was included in PD [53, 54, 75, 80, 86, 89, 122], tremor [133] and CP [12, 137], as well as the inter-quartile range for PD [82, 89]. The range of jerk and angular acceleration was used in one study in PD [72].

Peak-to-peak and magnitude of angular velocity were additionally used in PD [73, 77, 80, 87], whereas Repnik and colleagues calculated a rotational jerk index for angular velocity values to evaluate hand rotation in stroke [28]. Finally, a study in PD used the square root of the sum of squares of jerk signals and named this feature ‘segment velocity’ [48].

As basis statistical features, kurtosis and skewness were popular in PD [66, 76, 78, 86, 87, 118], but not in other populations apart from one ataxia study [134]. With respect to signal dynamics, multiple forms of entropy were used, most commonly sample entropy and approximate entropy in PD [53, 55, 62, 66, 70, 76, 86], stroke [120] and ataxia [136] and Shannon entropy and permutation entropy in PD [87], dyskinetic CP [139] and HD [29]. Fuzzy entropy was additionally calculated in two ataxia studies [131, 144]. Apart from entropy, the Gini index and Lyapunov exponent were additionally used as a measure of signal complexity in PD [87], ataxia [131] and HD [29]. The same HD study additionally used recurrence rate, determinism and average diagonal line to evaluate signal dynamics [29].

For signal smoothness, RMS of jerk was often used as a straightforward measure in PD [118] and stroke [115, 116], as well as a jerk metric for which multiple definitions were given, mostly RMS jerk normalized over time/peak velocity or mean jerk [12, 15, 76, 89, 90, 115, 123, 139, 141]. Additionally, smoothness measures were also described as the difference between movement accelerometer readings and smoothed readings, number of movement units or number of speed peaks [114, 122, 141].

Coefficient of variation was often used as a measure of variability or rhythm for different signals such as excursion angle [58, 69, 79, 101], (angular) velocity [58, 64, 79], amplitude [64, 71] and movement frequency [64, 83, 144], while two studies in PD defined ‘rhythm’ via the STD of intervals of a finger tapping movement [51] and any sequence of regularly occurring events [67]. Finally, a stroke study defined variability as the RMS error between a reference trial and a warped trial [117]. Considering the geometrical structure of a non-linear time-series, Newman et al. included Higuchu’s fractional dimension in children with CP [12].

With respect to orientation and rotational information, correlation between the different axes of the accelerometer or gyroscope was often included as a feature in PD [76, 86, 89, 99], HD [29] and stroke [115]. Additionally, the peak of the normalized cross-correlation from pairs of acceleration time series and the lag of first peak in autocorrelation acceleration were included in two PD studies [53, 56]. Concerning trajectories and travelled distances, multiple studies used different definitions for this feature. 3D hand trajectory and length of 3D trajectory [121] and path-length ratio were used in stroke [114], while the index of curvature (deviation from a straight line) was used in dyskinetic CP [13]. Elevation angle was included in a CP study, while in stroke, the similarity of hand trajectories was used [12, 28]. Two studies in patients with ataxia used mean and standard deviation of Euclidian distance from the mean trajectory and curved and straight-line similarity analysis [132, 135]. In PD, Heldman et al. used a bradykinesia index, based on variability in time and amplitude of task execution whereas Tamas et al. and Garza-Rodriguez and colleagues quantified hypokinesia using velocity decrement, which is defined as a decrease in velocity between subsequent data parts [69, 82].

In the frequency-domain, the dominant frequency component of acceleration/angular velocity or both was most often used [47, 53, 84, 87, 91, 94, 97-99, 104, 112, 115, 126, 127, 129], while only three studies included the second dominant frequency or dominant frequency of jerk [87, 115, 136]. Energy in the frequency spectrum was often included in multiple populations, both for the acceleration signal [49, 53, 56, 66, 115, 119, 120], angular velocity signal [100] or both [15, 62, 129]. One PD study additionally included amplitude and dominant frequency of modulations associated with the acceleration signal as well as fractal dimension [49], while one HD study included the average magnitude of the first five Short-Term-Fourier-Transfer components [29]. Apart from the frequency, power in specific frequency bands was a popular feature in multiple populations, including spectral power [68, 77, 84, 87, 102, 113, 118, 140], peak power [59, 60, 73, 87, 94], total power [59, 60, 73, 80, 99, 102, 104, 105, 110, 125, 134], mean power [63] and band power [139]. Considering entropy in the frequency domain, spectral entropy was used in two PD studies [62, 87] and one ataxia study [131], as well as component entropy in HD [29]. Spectral Arc Length was used as a measure of smoothness in two PD studies and one CP study [12, 73, 80]. For tremor studies, tremor frequency and tremor amplitude [109] were included as well as multiple specific tremor indices: Carpinella et al. defined the tremor index as the ratio of tremor (defined by peaks in the frequency spectrum) and the norm of angular velocity [124]. Western et al. defined average tremor amplitude as the product of frequency bins, sampling period and the signal’s power spectral density [128], whereas Benito-Leon et al. and McGurrin et al. used the mean logarithmic tremor power and tremor rotational amplitude/amplitude respectively, based on identified peaks in the power spectrum [103, 106].

### Statistical method used

Figure 4 gives a representation of the statistical methods used in the included studies. Forty-five studies included between-or within-group comparisons using statistical tests [12, 13, 28, 47, 51, 55, 58-60, 62-65, 69, 70, 73, 75, 79, 80, 85, 89, 90, 92-94, 96-98, 102, 104, 105, 109, 111, 114, 117, 123, 124, 131, 134, 137, 140-142, 144], mainly parametric and non-parametric ANOVA and parametric and non-parametric t-tests, whereas 43 studies used machine learning [15, 29, 48, 49, 53, 55-57, 62, 63, 66, 67, 72, 74-78, 81-84, 86-89, 100, 102, 112, 113, 115, 118-120, 122, 126, 129, 132, 133, 135, 136, 139, 144]. Twenty-five studies evaluated correlation of sensor features with clinical scales [12, 28, 51, 52, 54, 59, 60, 65, 70, 71, 80, 93, 103, 106-108, 110, 127-129, 131, 133, 134, 138, 144], whereas nine studies used regression analysis for a similar purpose [13, 54, 101, 102, 109, 116, 121, 133, 137]. Finally, five studies used only descriptive statistics or observation without hypothesis testing [11, 68, 91, 99, 125] and two studies evaluated ROC curves [61, 133]. The sum of these numbers does not add up to 101, because multiple studies used multiple of the abovementioned methods. Eleven studies used both statistical tests for comparison between groups and correlation with a clinical scale [12, 28, 59, 60, 65, 70, 71, 80, 93, 131, 134], whereas five studies used statistical tests and machine learning [55, 62, 63, 75, 89]. Yokoe et al. used both logistic regression and correlation with a clinical scale in PD [54]. In patients with ataxia, Tran et al. used statistical tests, correlation with a clinical scale and machine learning [144] and Oubre et al. used statistical tests, regression, correlation with a clinical scale and machine learning [133]. In participants with essential tremor, Ali et al. used statistical tests, regression analysis and machine learning [102] and Sprdlik et al. used statistical tests, ROC curves and regression analysis [109]. In CP, Sanger et al. used both regression analysis and ANOVA/t-tests [13], and Strohrmann et al. used a t-test, to subsequently continue with a linear regression for the features that were significantly different between groups [137].

**Figure 4:**
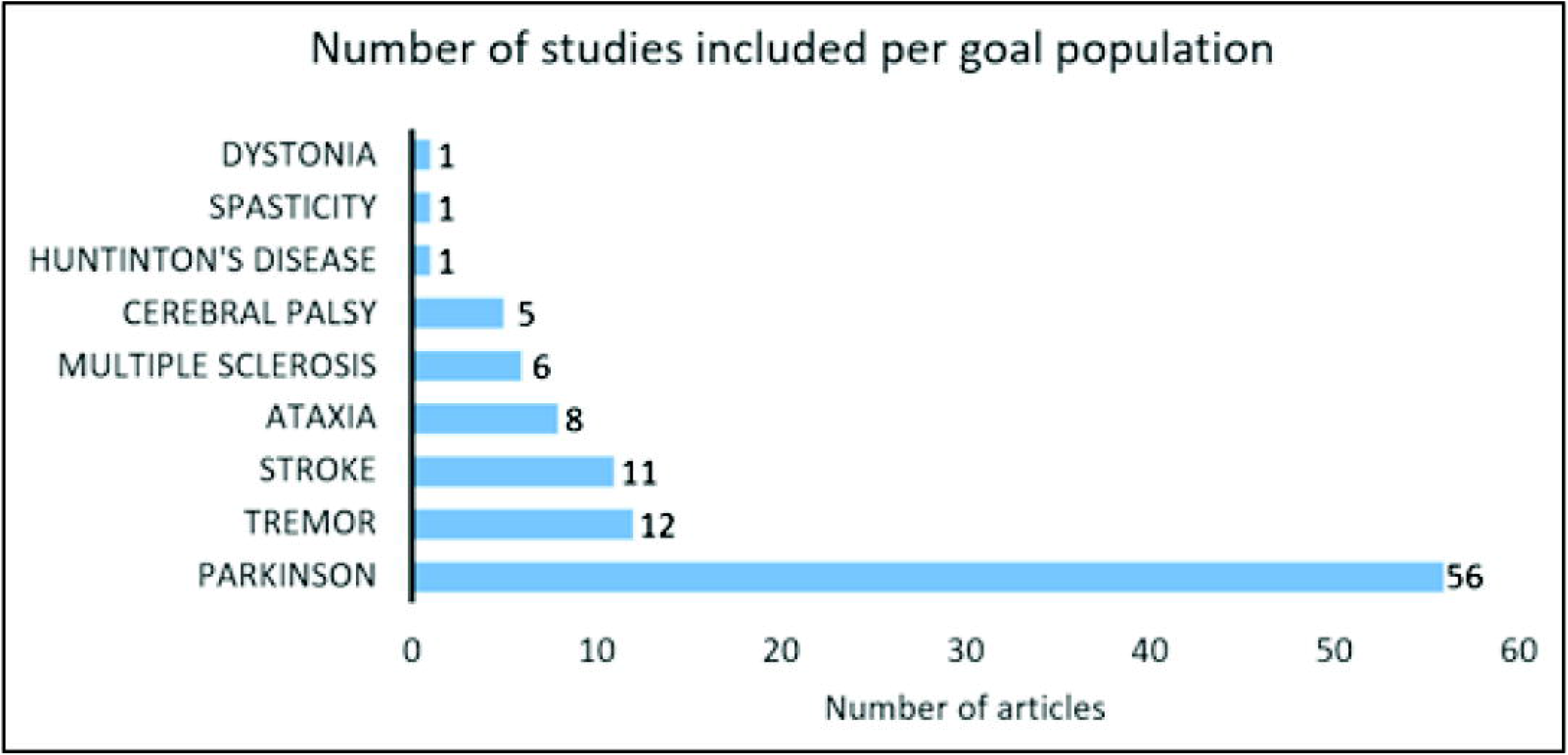
Statistical method used in the included studies. The sum does not add up to 101 because multiple studies used more than one methodology.

### Sensitivity and/or responsiveness of most prevalent sensor features

Table 3 provides an overview of the features included by more than five articles, the number of articles reporting sensitivity of the specific feature and the number of articles that identified a significant difference between groups, severity levels or pre/post intervention.

**Table 3:**
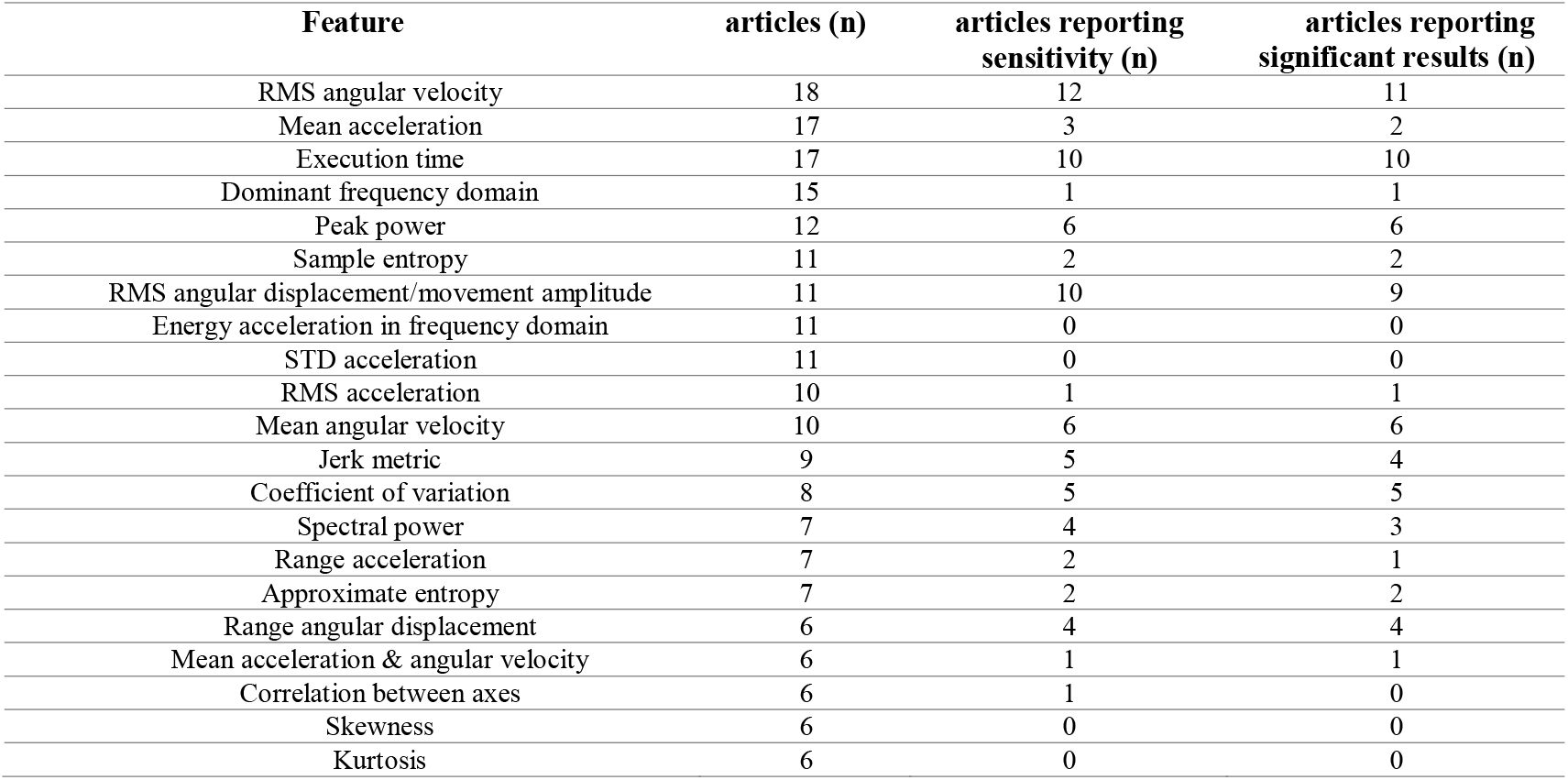
Most prevalent features, the number of articles they appear in, the number of articles reporting sensitivity and the number of articles reporting ignificant results.

| Feature | articles (n) | articles reporting sensitivity (n) | articles reporting significant results (n) |
| --- | --- | --- | --- |
| RMS angular velocity | 18 | 12 | 11 |
| Mean acceleration | 17 | 3 | 2 |
| Execution time | 17 | 10 | 10 |
| Dominant frequency domain | 15 | 1 | 1 |
| Peak power | 12 | 6 | 6 |
| Sample entropy | 11 | 2 | 2 |
| RMS angular displacement/movement amplitude | 11 | 10 | 9 |
| Energy acceleration in frequency domain | 11 | 0 | 0 |
| STD acceleration | 11 | 0 | 0 |
| RMS acceleration | 10 | 1 | 1 |
| Mean angular velocity | 10 | 6 | 6 |
| Jerk metric | 9 | 5 | 4 |
| Coefficient of variation | 8 | 5 | 5 |
| Spectral power | 7 | 4 | 3 |
| Range acceleration | 7 | 2 | 1 |
| Approximate entropy | 7 | 2 | 2 |
| Range angular displacement | 6 | 4 | 4 |
| Mean acceleration & angular velocity | 6 | 1 | 1 |
| Correlation between axes | 6 | 1 | 0 |
| Skewness | 6 | 0 | 0 |
| Kurtosis | 6 | 0 | 0 |

RMS of angular velocity was reported in 18 studies, with sensitivity results for 11 studies. In PD, Van den Noort et al. found significantly higher RMS values for ON vs OFF dopaminergic medication, while Summa et al. did not find a significant difference between medication states [11, 73]. Espay et al. found 25% improvement in RMS values after dopaminergic medication in PD patients [58]. Kwon et al. and Luksys et al. found significant higher RMS angular velocity values for PD patients in comparison with controls, whereas two studies found a correlation of -0.78 between RMS angular velocity and clinical scores of the UPDRS [59, 60, 79, 98]. Additionally, Heldman et al. found a correlation of -0.78 between RMS angular velocity values and the modified bradykinesia rating scale [101] and Salarian et al. found good correlation between RMS angular velocity values and the UPDRS bradykinesia subscore, as well as good correlation between RMS angular velocity of the roll axis and the tremor subscore of the UPDRS [52]. In patients with tremor, spearman correlation between RMS angular velocity and tremor severity scores ranged from 0.19 (finger-to-nose) to 0.73 (keeping arms extended in front of the body) for Lopez-Bianco et al. [108] and between 0.41 and 0.70 for Kwon et al. [110], whereas Heo et al. found lower RMS angular velocity values after electrical stimulation [105].

Seventeen studies reported mean acceleration as a feature, but only two PD studies and one ataxia study discussed its sensitivity. Romano et al. found lower mean acceleration for PD patients in comparison with the control group, while Zwartjes et al. did not find significant differences between ON and OFF stimulation states of deep brain stimulation [57, 90]. In patients with Ataxia, Samotus et al. found lower mean acceleration after botulinum-toxin-A injections [111]. Execution time was included in 17 studies, with reported sensitivity for 11 studies. Execution time significantly differed between different severity levels [28] and between healthy controls and patients with stroke [28, 117] and MS [123, 124] and between the paretic and non-paretic arm in children with unilateral CP [12]. Execution time was significantly longer for PD patients in comparison with healthy controls [75, 80] and for patients with multiple system atrophy of parkinsonian type and progressive supranuclear palsy in comparison with healthy controls [71]. Third, execution time was significantly different between the ON and OFF medication state in PD patients [11]. In CP, execution time was one of the three features to best estimate upper limb performance in a regression analysis [137].

The dominant frequency domain was included in 15 studies, but only Hoff et al. reported individual contributions of this feature, reporting that amplitude in 1-4Hz and 4-8Hz frequency bands correlated with the modified Abnormal Involuntary Movement Scale [47]. Peak power was included in 12 studies, of which six discussed its individual sensitivity. Jun et al. reported a good correlation between peak power and clinical bradykinesia scores and Kim et al. reported decreasing peak powers with increasing UPDRS scales steps [59, 60], while Makabe et al. reported increasing peak powers with increasing severity stages of the Hoehn and Yahr scale [94]. Similarly, Summa et al. reported increases in peak power in ON vs OFF medication state [73]. In essential tremor, Heo et al. reported higher peak power after electrical stimulation [105] and Kwon et al. reported high correlation between peak power and tremor severity scores [110].

Sample entropy was included in 11 studies, but only two PD studies reported its sensitivity. Chelaru et al. found significantly higher entropy for dyskinetic PD patients in comparison with non-dyskinetic PD patients, as well as Liu et al. who found a significant difference between PD patients and healthy controls and good correlation with UPDRS scores [55, 70]. RMS of angular displacement was included in 11 studies, of which ten reported sensitivity. Tamas et al. found significant differences in RMS amplitude before and after subthalamic stimulation and Espay et al. found significant differences between ON and OFF medication state in PD [58, 69]. Kwon et al. found significantly lower RMS amplitudes for PD patients in comparison with controls and Jun et al. found decreasing angular displacement with increasing bradykinesia scores, but this was based on visual observation [60, 79]. Chan et al. found higher values for angular displacement for patients with PD with tremor in comparison with essential tremor [92]. Kim et al. additionally found a significant difference between PD patients and controls [59], whereas Heldman et al. found a correlation of -0.81 between RMS excursion angle and clinical scores [101]. Delrobaei et al. found a higher tremor severity score (which was composed of the RMS values of angular velocity) for tremor-dominant PD patients in comparison with non-tremor dominant PD patients and good correlation between tremor severity score and UPDRS scores [93]. In essential tremor, Kwon et al. and Chan et al. found correlations ranging from 0.29-0.66 and 0.80-0.93 respectively, between RMS angular displacement and tremor severity scores [107, 110]. Energy and STD of acceleration were included in 10 studies, but none reported sensitivity.

RMS of acceleration was included in 10 studies, but only van den Noort et al. discussed its specific contribution in PD patients, reporting increased RMS acceleration in ON vs OFF medication state during a finger tapping and opening/closing of the hand task [11]. Mean angular velocity was also included in 10 studies with six of them reporting sensitivity. In PD, three studies found lower mean angular velocity for PD patients in comparison with healthy controls [64, 71, 90], whereas one study additionally identified significant differences between ON/OFF DBS stimulation [52]. Garza-Rodriguez et al. found lower angular velocity values for PD patients with higher clinical severity [81]. In patients with ataxia, Oubre et al. found significant differences between patients and healthy controls [133].

Jerk metrics were calculated in nine studies with five reporting on its sensitivity. Romano et al. used the dimensionless jerk index as a jerk metric and found a significant difference between PD patients and healthy controls, while Habets et al. did not find a significant difference between ON and OFF medication state in PD patients [89, 90]. Carpinella et al. found a significantly higher jerk measure for patients with MS in comparison with healthy controls and a negative correlation between the jerk measure and ARAT score (r = -0.90) [123]. In children with unilateral CP, Newman and colleagues found a significantly higher normalised jerk index for the paretic arm in comparison with the non-paretic arm, but no correlation with the Melbourne Assessment Scale [12]. In children with spasticity, the normalized jerk score improved significantly after botulinum-toxin A injections [141].

Coefficient of variation (CoV) was included in eight studies, where CoV of time and amplitude was mostly calculated to evaluate bradykinesia. Djuric-Jovicic and colleagues found significant differences between PD patients and healthy controls for both CoV of time and amplitude, whereas Lee et al. found significant differences for CoV of speed, amplitude and frequency between PD patients and controls [64, 71]. Kwon et al. additionally found significant differences between PD patients and controls for the CoV of angles and velocity [79]. Tamas et al. found that the coefficient of variation – also called ‘rhythm’ – improved significantly after bilateral and contralateral subthalamic stimulation, whereas Espay et al. found significant differences between ON and OFF medication state for CoV in PD patients [58, 69].

Spectral power was used in seven studies of which four reported sensitivity. Bravo et al. compared power spectral density (PSD) plots between PD patients and healthy controls and found both higher and lower PSD amplitude for PD patients in comparison with healthy controls, depending on the individual [68]. In patients with dystonia, Legros et al. found a decrease of the area under the spectrum curve after deep brain stimulation surgery [140]. Ali et al. found higher PSD ratios for patients with essential tremor in comparison with healthy controls [102], whereas Heldman et al. found correlations from 0.77-0.83 between the logarithm of peak power and the UPDRS scores [113]. The range of acceleration was additionally calculated in seven articles, but only two articles reported its sensitivity. Rabelo et al. found a significantly higher acceleration range for healthy controls in comparison with PD patients, while Habets et al. did not find a significant difference between ON and OFF medication state in PD patients [75, 89]. Approximate entropy was also included in seven studies, but only two PD studies included its sensitivity, where Liu et al. and Luksys et al. found significant differences between PD patients and a control group [70, 98].

Range of angular displacement was calculated in six studies, but only four discussed its sensitivity. Djuric et al. reported a higher range for healthy controls in comparison with PD patients, whereas van den Noort et al. reported lower displacement in the ON vs OFF medication state and improved amplitude in the ON compared to OFF state [11, 71]. Romano et al. found significant differences between PD patients and healthy controls for wrist flexion and shoulder movements and Salarian et al. found significantly lower angular displacements at the level of the wrist for PD patients compared to healthy controls [52, 90]. Energy of acceleration in the frequency domain and STD of acceleration were included in 11 articles, but all of them included these features as part of a feature set for machine learning, without discussing its individual contribution.

Mean acceleration and angular velocity were included in six studies, but only Romano et al. found significantly lower mean acceleration and angular velocity in PD patients in comparison with healthy controls [90]. Correlation between axes was included in six studies, but only Zhu et al. reported no significant differences in correlations when comparing PD patients in ON and OFF medication state [99]. Kurtosis and skewness were additionally included in six studies, but none of them reported the contribution of the individual features.

### Clinical application

Figure 5 presents an overview of the clinical application of the included studies. Fifty-two studies used sensor features for the automatization of a clinical scale [15, 29, 49, 51-54, 56, 57, 59, 60, 63, 65, 67, 72, 76-78, 81-84, 86-88, 93, 100, 101, 103, 106-108, 110, 112, 113, 115, 116, 118-123, 126-129, 131, 133, 137-139]. Sixteen studies used sensor features to evaluate the effect of an intervention [47, 48, 50, 55, 58, 68, 69, 74, 89, 99, 104, 105, 111, 140-142], whereas 35 studies used the obtained features to discriminate between patients and controls or between different patient groups [11-13, 28, 61, 62, 64-66, 70-73, 75, 79, 80, 85, 90-94, 96-98, 102, 109, 117, 124, 125, 132, 134-136, 144]. Four studies subsequently discriminated between different severity levels [28, 65, 72, 114]. Again, there was some overlap in clinical applications: Delrobaei et al., Spasojevic et al. and Repnik et al. compared control and patient groups as well as severity levels within the patient group, while also correlating sensor features with a clinical scale [28, 65, 72]. Kamper et al. compared a patient and control group but also compared severity levels separately [114].

**Figure 5:**
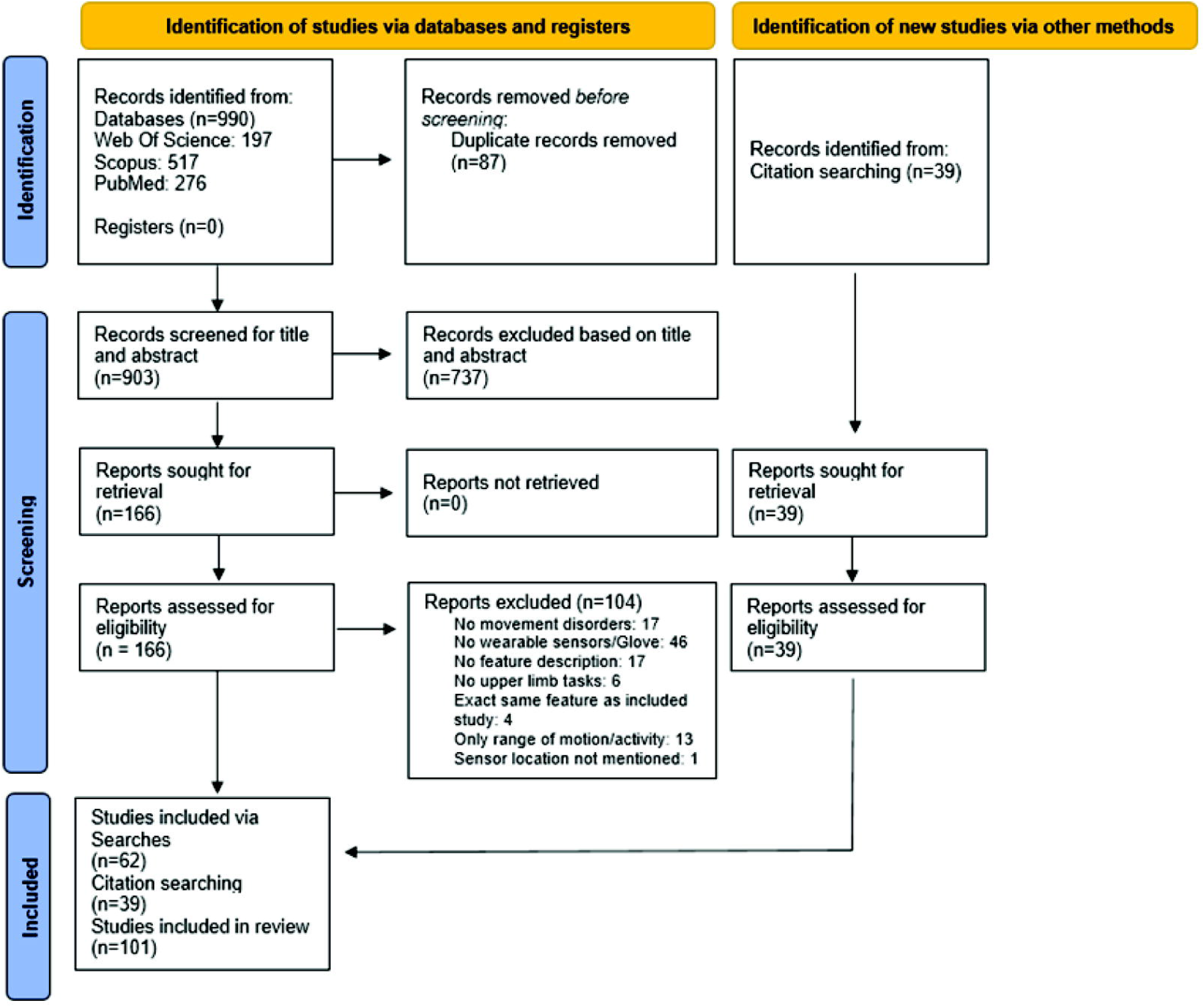
Clinical application. The sum does not add up to 101 because multiple studies used more than one methodology.

## 4 Discussion

The primary objective of this systematic review was to provide an overview of sensor set-up and type, included tasks, sensor features and statistical methods that are used to evaluate movement disorders during upper limb tasks in multiple pathological populations. We identified 101 studies in eight pathological conditions using wearable sensors placed on the upper limb during upper limb tasks and including at least one sensor feature based on linear acceleration or angular velocity. Of all included studies, 55% were studies in PD, 12% were studies with essential tremor patients, 11% were studies in stroke patients, 8% were studies in adults or children with ataxia, 6% were studies including participants with MS and 5% included children with CP. Adults with HD and spasticity and dystonia in children represented only 1% of the included studies. When comparing these numbers with the prevalence of the abovementioned conditions, an important imbalance emerges. Worldwide, approximately 101 million people are living post-stroke [147], 25 million people live with essential tremor [148], 17 million people live with CP [149], 10 million people are estimated to live with PD [150-152], approximately 0.2 to 3 million people live with ataxia, depending on the type [153, 154] and 0.2 to 0.5 million people live with HD, depending on the geographical area [155, 156]. While stroke is much more prevalent than PD or essential tremor, this ratio is not reflected in the number of available studies per condition. More surprisingly, where CP is the most prevalent neurological childhood condition included apart from stroke and essential tremor, its high prevalence does not agree with the number of studies investigating the associated movement disorders using wearable sensors. Current findings thus identify a mayor gap between prevalence of a condition and insights in the related movement disorders. Especially for early-onset conditions such as CP, more insights in the disturbed movement patterns from an early age could benefit targeted therapy and long-term treatment management.

The abundance of included PD studies reflects its more advanced state-of-the-art assessment in comparison with other pathological populations. These insights offer opportunities and learning experiences for clinicians and researchers aiming to bridge the gap between technology and clinical measures in the quantitative evaluation of movement disorders. Although wide-spread in research, the clinical implementation of IMU-based analysis of movement disorders is lacking in clinical practice in all populations, mainly due to the lack of validation of algorithms in real-world conditions [157].

With respect to sensor type, IMUs containing both an accelerometer and gyroscope were most often used, where a time-related trend was clearly visible in the included PD studies: between 2000 and 2010, all PD studies included either an accelerometer or a gyroscope, whereas after 2010, IMUs were almost exclusively used. This trend is presumably supported by technological advancements, allowing more sensors in a smaller device with longer battery life combined with more affordable prices for IMUs.

Sensor location, number of included sensors and upper limb tasks were separately discussed to provide a comprehensive overview, but conclusions should be drawn on a combination of these settings as they are closely inter-related. E.g., five out of the ten studies that placed one sensor on the index finger included the finger tapping task [51, 59, 61, 69, 113] and of the nine studies who placed a sensor one the thumb and index finger, all included finger tapping [54, 58, 64, 70, 71, 73, 85, 88, 101]. When more sensors were used both on the distal and proximal upper limb, tasks ranged from distal upper limb tasks (pro/supination) [65, 74] to a myriad of tasks such as the ARAT [28]. Finally, the four studies that measured activities in a home environment all placed sensors on the wrist, mostly likely due to the high comfort and ease of use of wrist-worn sensors [63, 89, 134, 139].

The collection of upper limb tasks included in the selected studies reflects the insight that the choice of upper limb task is heavily dependent on the movement disorder. The high prevalence of finger tapping and wrist pro/supination in the PD studies follows from their presence in the Motor Examination part of the (MDS-)UPDRS [20], whereas the finger-to-nose task and keeping arms extended in front of the body are part of both the (MDS-)UPDRS and the Essential Tremor Rating Assessment Scale [22]. Both tasks are well-suited to quantify decrease and slowness of movements. Since these scales are well implemented in clinical practice, patients are often requested to perform these tasks in the presence of a neurologist. In stroke, the Wolf Motor Function Task was most popular, presumably because this scale is used in daily practice for the evaluation of upper extremity rehabilitation progress. An important notion is that the aetiological differences between PD/tremor on the one hand and CP, stroke and dystonia on the other hand influence the potential of task execution. In CP and stroke, functional ability can be impaired to a level where execution of specific functional tasks is not possible, which requires a very different approach in comparison with PD or tremor, where most tasks can be executed but performance may be impaired. When the level of physical impairment prohibits the execution of specific tasks, one should focus on monitoring of the movement disorders during home-based activities such as powered mobility (e.g. joy-stick steering) or in rest positions in the case of severe CP or stroke [139].

The secondary objective was to identify the most sensitive sensor features for symptom detection and quantification and describe the application of the proposed methods in clinical practice. Similar to the requested tasks, the derived sensor features were dependent of the movement disorder under investigation. Mean amplitude, movement/amplitude decrement and RMS, range and IQR of angular displacement were only used in PD studies and are hypothesized to correlate with the definition of hypokinesia (reduction in movement amplitude) in the (MDS-)UPDRS. Range and RMS of angular displacement can detect differences between PD and TD groups and quantify the severity of hypokinesia, implying that these features can be used in clinical practice as simply interpretable triggers of movement reduction. Velocity decrement and peak-to-peak, magnitude, IQR and mean of angular velocity were additionally only used in PD studies and are hypothesized to relate to the bradykinesia (slowing of movement) aspect in the (MDS-)UPDRS, emphasizing their clinical usefulness for early detection of bradykinesia symptoms [81]. Coefficient of variation of both amplitude and velocity as well as rhythm, were included to reflect the interruptions as described in the (MDS-)UPDRS. CoV values are easy to calculate and interpret and showed to be sufficiently sensitive to discriminate between medication and stimulation states in PD patients from both finger- and wrist-worn sensors. This parameter could thus be implemented to evaluate objective intervention effects in large-scale medication or stimulation studies. In essential tremor and studies focusing on tremor in PD patients, occurrence and amplitude of peaks in specific frequency bands as well as power in these frequency bands were most often included, owing to the rhythmical aspect of tremor. However, the selected frequency bands were not always similar. The 4-12Hz frequency band was most often considered as tremor [102, 113], while Heo et al. and Kwon et al. used 3-12Hz [105, 110], Patel et al. used a 3-8Hz band and Schaefer et al. considered 7-12Hz as the tremor frequency [53, 96]. Makabe et al. used a range of 8-12Hz and 20-25Hz and Sprdlik et al. used the frequency distribution for highest maxima between 1-15Hz [94, 109]. Lopez-Bianco et al. used a high-pass filter with cut-off 4Hz followed by low-pass filter with cut-off 8Hz [108]. These differences suggest that a solid definition of tremor frequency is required in order to standardize instrumented tremor quantification, to allow comparison of methodologies on a large-scale cross-sectional level and to facilitate data merging and sharing.

In pathologies not related to PD or tremor, path length or similarity of hand trajectories were calculated. This was the case in stroke, dyskinetic CP and spasticity, reflecting the importance of the impact of the movement disorder on reaching movements. The frequent use of sensor features such as smoothness and jerk metrics might reflect the effect of the location of the brain lesions on the smooth execution of functional tasks and its impact on daily-life activities in these pathologies.

The clinical application of the included studies varied from discrimination of groups to prediction of severity levels and was closely related to the method used to obtain this specific result. With respect to the discrimination of groups, the sensor features sufficiently sensitive to detect differences between a control group and pathological patients could be used for early detection of e.g. PD or MS symptoms, allowing for early intervention and possibly preventing rapid worsening of symptoms. For the prediction of severity levels, all PD studies correlated the sensor features to the (MDS-)UPDRS (or specific sub-items), the AIMS or the Hoehn & Yahr scale. In CP and stroke, sensor features were correlated with the Melbourne Assessment Scale and ARAT, whereas in another CP study, the Jebsen-Taylor Test, the Quality of Upper Extremity Skills Test (QUEST) and the Box and Blocks Test were included. When the clinical application was the (side)effect of intervention, six out of 16 studies used sensor features to assess dyskinesia in PD patients, as this is a well-known levodopa-induced motor complication [158]. The (MDS-)UPDRS, Hoehn & Yahr and Essential Tremor Rating Assessment Scale are severity scales, while the Melbourne Assessment Scale, the ARAT, the Jebsen-Taylor Test, the Quality of Upper Extremity Skills Test and the Box and Blocks Test evaluate upper extremity function. In stroke and CP however, the severity of the movement disorder is dependent on the location of the brain lesion, which was not studied in detail in the included studies and has not been fully elucidated to date in most movement disorders. To this end, wearable sensors provide opportunities for detailed exploration of the connection between the location of the brain lesion and the aetiology and severity of movement disorders.

IMUs have mostly been used to assess upper limb use and for detection of activity periods in daily life in patients with PD and/or essential tremor [159-161], CP [162-164] or stroke [165], but their application to quantify movement disorders in the upper limb is less extensive. Activity measures mostly focus on the amount of time that acceleration measures exceed a pre-defined threshold (e.g., Activity Index), which yields information about the quantity of movement, but not about the quality. To facilitate follow-up of intervention or long-term rehabilitation programs, a combined assessment of both movement quantity and quality can provide more insights in both the presence and severity of movement disorders. Ideally, long-term monitoring is executed in a home-environment (i.e., low patient-burden while collecting long-term data), while a contact moment to record pathology-related tasks in a standardized setting could be added to the study protocol since this allows more specific data analysis, e.g., through the presence of video recordings of the performed tasks.

The lessons learned from the PD studies in the current review can accelerate the development of wearable sensors protocols in the remaining pathologies, provided that there is sufficient attention for the standardisation of protocols, tasks, feasibility and data analysis methods. These conditions could facilitate reproduction of studies, large-scale multi-centre studies and merging of study results in the near future. The insights provided in current review highlight the potential of wearable sensors for symptom detection and evaluation in CP, stroke, HD, MS and dystonia, but a larger international research focus is urgently required to meet those needs.

### Conclusions and future directions

Wearable sensors offer a myriad of opportunities for the quantification of movement disorders in multiple pathologies, but the abundance of available information could threaten its usability. Our findings illustrate that there are a lot of similarities between pathology-related sensor protocols and tasks, but the agreement is yet not sufficient to allow data pooling or international multi-centre studies. For this purpose, higher-level standardisation with respect to task selection and sensor feature extraction per pathology is strongly recommended. Although multiple sensors can provide a lot of information, researchers should think carefully about the balance between information gain and accessibility. One sensor on the index finger for PD or on the wrist for other pathologies could be attached in a non-obstructive way, allowing for better adherence and less missing data due to e.g., battery loss. Current overview can support clinicians and researchers to select the most sensitive pathology-dependent sensor features and measurement methodologies for detection and quantification of upper limb movement disorders and for the objective evaluations of treatment effects.

## Supporting information

PRISMA checklist

## Data Availability

All data produced in the present study are available upon reasonable request to the authors

## 5 Author Contributions

**IV**: Conceptualisation, methodology, analysis, data curation, writing-original draft, visualisation. **HH**: Methodology, writing-reviewing and editing. **JDV**: Writing-reviewing and editing. **EVW:** Reviewing and editing. **HF**: Writing-reviewing and editing. **KD**: Writing-reviewing and editing. **J-MA**: Writing-reviewing and editing. **EM:** Conceptualisation, writing-reviewing and editing, supervision, project administration, funding acquisition.

## 7 Acknowledgments

The author would like to thank M.N. and S.B. for their help throughout the systematic review process.

## 8 Supplementary Material

Supplementary Material should be uploaded separately on submission, if there are Supplementary Figures, please include the caption in the same file as the figure. Supplementary Material templates can be found in the Frontiers Word Templates file.

Please see the <u>Supplementary Material section of the Author guidelines</u> for details on the different file types accepted.

## 9 Data Availability Statement

The data that support the findings of this study are available from the corresponding author upon reasonable request.

## References

1. Kukke, S.N., et al., Coordination of Reach-to-Grasp Kinematics in Individuals With Childhood-Onset Dystonia Due to Hemiplegic Cerebral Palsy. IEEE Trans Neural Syst Rehabil Eng, 2016. 24(5): p. 582–590.

2. Levin, M.F., Interjoint coordination during pointing movements is disrupted in spastic hemiparesis. Brain, 1996. 119(1): p. 281–293.

3. Beer, R.F., J.P.A. Dewald, and W.Z. Rymer, Deficits in the coordination of multijoint arm movements in patients with hemiparesis: evidence for disturbed control of limb dynamics. Experimental Brain Research, 2000. 131(3): p. 305–319.

4. Jankovic, J., et al., Chapter 1 - Clinical overview and phenomenology of movement disorders, in Principles and Practice of Movement Disorders (Third Edition), J. Jankovic, et al., Editors. 2021, Elsevier: London. p. 1–51.e27.

5. Wenning, G.K., et al., Prevalence of movement disorders in men and women aged 50-89 years (Bruneck Study cohort): a population-based study. Lancet Neurol, 2005. 4(12): p. 815–20.

6. Monbaliu, E., et al., Clinical presentation and management of dyskinetic cerebral palsy. The Lancet Neurology, 2017. 16(9): p. 741–749.

7. Nutt, J.G., et al., Epidemiology of focal and generalized dystonia in Rochester, Minnesota. Mov Disord, 1988. 3(3): p. 188–94.

8. Phukan, J., et al., Primary dystonia and dystonia-plus syndromes: clinical characteristics, diagnosis, and pathogenesis. The Lancet Neurology, 2011. 10(12): p. 1074–1085.

9. Müller, J., et al., The prevalence of primary dystonia in the general community. Neurology, 2002. 59(6): p. 941–943.

10. Tysnes, O.B. and A. Storstein, Epidemiology of Parkinson’s disease. J Neural Transm (Vienna), 2017. 124(8): p. 901–905.

11. van den Noort, J.C., et al., Quantification of Hand Motor Symptoms in Parkinson’s Disease: A Proof-of-Principle Study Using Inertial and Force Sensors. Ann Biomed Eng, 2017. 45(10): p. 2423–2436.

12. Newman, C.J., et al., Measuring upper limb function in children with hemiparesis with 3D inertial sensors. Childs Nerv Syst, 2017. 33(12): p. 2159–2168.

13. Sanger, T.D., Arm trajectories in dyskinetic cerebral palsy have increased random variability. J Child Neurol, 2006. 21(7): p. 551–7.

14. Lee, K.B., et al., Six-month functional recovery of stroke patients: a multi-time-point study. International journal of rehabilitation research. Internationale Zeitschrift fur Rehabilitationsforschung. Revue internationale de recherches de readaptation, 2015. 38(2): p. 173–180.

15. Zhang, M., et al., Beyond the standard clinical rating scales: fine-grained assessment of post-stroke motor functionality using wearable inertial sensors. Annu Int Conf IEEE Eng Med Biol Soc, 2012. 2012: p. 6111–5.

16. Cohen, N., et al., Multidisciplinary intensive outpatient rehabilitation program for patients with moderate-to-advanced Parkinson’s disease. NeuroRehabilitation, 2021. 49(1): p. 47–55.

17. Jackman, M., et al., Predicting Improvement in Writer’s Cramp Symptoms following Botulinum Neurotoxin Injection Therapy. Tremor Other Hyperkinet Mov (N Y), 2016. 6: p. 410.

18. Umar, M., T. Masood, and M. Badshah, Effect of botulinum toxin A & task-specific training on upper limb function in post-stroke focal dystonia. J Pak Med Assoc, 2018. 68(4): p. 526–531.

19. Movement Disorder Society Task Force on Rating Scales for Parkinson’s Disease, The Unified Parkinson’s Disease Rating Scale (UPDRS): Status and recommendations. Movement Disorders, 2003. 18(7): p. 738–750.

20. Goetz, C.G., et al., Movement Disorder Society-sponsored revision of the Unified Parkinson’s Disease Rating Scale (MDS-UPDRS): scale presentation and clinimetric testing results. Mov Disord, 2008. 23(15): p. 2129–70.

21. Hoehn, M.M. and M.D. Yahr, Parkinsonism: onset, progression, and mortality. 1967. Neurology, 1998. 50(2): p. 318 and 16 pages following.

22. disease, M.D.S.T.F.o.R.S.f.P.s., Reliability of a new scale for essential tremor. Movement disorders : official journal of the Movement Disorder Society, 2012. 27(12): p. 1567–1569.

23. Schmitz-Hübsch, T., et al., Scale for the assessment and rating of ataxia: development of a new clinical scale. Neurology, 2006. 66(11): p. 1717–20.

24. Fugl-Meyer, A.R., et al., The post-stroke hemiplegic patient. 1. a method for evaluation of physical performance. Scand J Rehabil Med, 1975. 7(1): p. 13–31.

25. Wolf, S.L., et al., Assessing Wolf motor function test as outcome measure for research in patients after stroke. Stroke, 2001. 32(7): p. 1635–9.

26. Gladstone, D.J., C.J. Danells, and S.E. Black, The Fugl-Meyer Assessment of Motor Recovery after Stroke: A Critical Review of Its Measurement Properties. Neurorehabilitation and Neural Repair, 2002. 16(3): p. 232–240.

27. Platz, T., et al., Reliability and validity of arm function assessment with standardized guidelines for the Fugl-Meyer Test, Action Research Arm Test and Box and Block Test: a multicentre study. Clin Rehabil, 2005. 19(4): p. 404–11.

28. Repnik, E., et al., Using Inertial Measurement Units and Electromyography to Quantify Movement during Action Research Arm Test Execution. Sensors (Basel), 2018. 18(9).

29. Bennasar, M., et al., Automated Assessment of Movement Impairment in Huntington’s Disease. IEEE Trans Neural Syst Rehabil Eng, 2018. 26(10): p. 2062–2069.

30. Gilmore, R., L. Sakzewski, and R. Boyd, Upper limb activity measures for 5-to 16-year-old children with congenital hemiplegia: a systematic review. Dev Med Child Neurol, 2010. 52(1): p. 14–21.

31. Spirtos, M., P. O’Mahony, and J. Malone, Interrater reliability of the Melbourne Assessment of Unilateral Upper Limb Function for children with hemiplegic cerebral palsy. Am J Occup Ther, 2011. 65(4): p. 378–83.

32. Burke, R.E., et al., Validity and reliability of a rating scale for the primary torsion dystonias. Neurology, 1985. 35(1): p. 73–7.

33. Monbaliu, E., et al., The Dyskinesia Impairment Scale: a new instrument to measure dystonia and choreoathetosis in dyskinetic cerebral palsy. Developmental Medicine & Child Neurology, 2012. 54(3): p. 278–283.

34. Pang, Y., et al., Automatic detection and quantification of hand movements toward development of an objective assessment of tremor and bradykinesia in Parkinson’s disease. Journal of Neuroscience Methods, 2020. 333: p. 108576.

35. Agostino, R., et al., Impairment of individual finger movements in Parkinson’s disease. Movement Disorders, 2003. 18(5): p. 560–565.

36. Kreulen, M., et al., Movement patterns of the upper extremity and trunk before and after corrective surgery of impaired forearm rotation in patients with cerebral palsy. Dev Med Child Neurol, 2006. 48(6): p. 436–41.

37. Butler, E.E. and J. Rose, The pediatric upper limb motion index and a temporal-spatial logistic regression: quantitative analysis of upper limb movement disorders during the Reach & Grasp Cycle. J Biomech, 2012. 45(6): p. 945–51.

38. Simon-Martinez, C., et al., Effects of combining constraint-induced movement therapy and action-observation training on upper limb kinematics in children with unilateral cerebral palsy: a randomized controlled trial. Scientific Reports, 2020. 10(1): p. 10421.

39. Cuesta-Gómez, A., et al., Functional electrical stimulation improves reaching movement in the shoulder and elbow muscles of stroke patients: A three-dimensional motion analysis. Restor Neurol Neurosci, 2019. 37(3): p. 231–238.

40. Alt Murphy, M., et al., Kinematic Analysis Using 3D Motion Capture of Drinking Task in People With and Without Upper-extremity Impairments. J Vis Exp, 2018(133).

41. Lang, C.E., S.L. DeJong, and J.A. Beebe, Recovery of thumb and finger extension and its relation to grasp performance after stroke. J Neurophysiol, 2009. 102(1): p. 451–9.

42. Maetzler, W., et al., Quantitative wearable sensors for objective assessment of Parkinson’s disease. Mov Disord, 2013. 28(12): p. 1628–37.

43. Tortelli, R., F.B. Rodrigues, and E.J. Wild, The use of wearable/portable digital sensors in Huntington’s disease: A systematic review. Parkinsonism & Related Disorders, 2021. 83: p. 93–104.

44. Haberfehlner, H., et al., Instrumented assessment of motor function in dyskinetic cerebral palsy: a systematic review. Journal of NeuroEngineering and Rehabilitation, 2020. 17(1): p. 39.

45. Page, M.J., et al., The PRISMA 2020 statement: an updated guideline for reporting systematic reviews. BMJ, 2021. 372: p. 71.

46. Morgan, R.L., et al., Identifying the PECO: A framework for formulating good questions to explore the association of environmental and other exposures with health outcomes. Environment international, 2018. 121(Pt 1): p. 1027–1031.

47. Hoff, J.I., et al., Accelerometric assessment of levodopa-induced dyskinesias in Parkinson’s disease. Mov Disord, 2001. 16(1): p. 58–61.

48. Keijsers, N.L., M.W. Horstink, and S.C. Gielen, Automatic assessment of levodopa-induced dyskinesias in daily life by neural networks. Mov Disord, 2003. 18(1): p. 70–80.

49. Bonato, P., et al., Data mining techniques to detect motor fluctuations in Parkinson’s disease. Conf Proc IEEE Eng Med Biol Soc, 2004. 2004: p. 4766–9.

50. Koop, M.M., et al., Improvement in a quantitative measure of bradykinesia after microelectrode recording in patients with Parkinson’s disease during deep brain stimulation surgery. Movement Disorders, 2006. 21(5): p. 673–678.

51. Okuno, R., et al., Finger taps movement acceleration measurement system for quantitative diagnosis of Parkinson’s disease. Conf Proc IEEE Eng Med Biol Soc, 2006. Suppl: p. 6623–6.

52. Salarian, A., et al., Quantification of tremor and bradykinesia in Parkinson’s disease using a novel ambulatory monitoring system. IEEE Trans Biomed Eng, 2007. 54(2): p. 313–22.

53. Patel, S., et al., Monitoring Motor Fluctuations in Patients With Parkinson’s Disease Using Wearable Sensors. IEEE Transactions on Information Technology in Biomedicine, 2009. 13(6): p. 864–873.

54. Yokoe, M., et al., Opening velocity, a novel parameter, for finger tapping test in patients with Parkinson’s disease. Parkinsonism Relat Disord, 2009. 15(6): p. 440–4.

55. Chelaru, M.I., C. Duval, and M. Jog, Levodopa-induced dyskinesias detection based on the complexity of involuntary movements. Journal of Neuroscience Methods, 2010. 186(1): p. 81–89.

56. Cole, B.T., et al., Dynamic neural network detection of tremor and dyskinesia from wearable sensor data. Annu Int Conf IEEE Eng Med Biol Soc, 2010. 2010: p. 6062–5.

57. Zwartjes, D.G., et al., Ambulatory monitoring of activities and motor symptoms in Parkinson’s disease. IEEE Trans Biomed Eng, 2010. 57(11).

58. Espay, A.J., et al., Differential response of speed, amplitude, and rhythm to dopaminergic medications in Parkinson’s disease. Mov Disord, 2011. 26(14): p. 2504–8.

59. Kim, J.W., et al., Quantification of bradykinesia during clinical finger taps using a gyrosensor in patients with Parkinson’s disease. Med Biol Eng Comput, 2011. 49(3): p. 365–71.

60. Jun, J.-H., et al., Quantification of limb bradykinesia in patients with Parkinson’s disease using a gyrosensor — Improvement and validation. International Journal of Precision Engineering and Manufacturing, 2011. 12(3): p. 557–563.

61. Hoffman, J.D. and J. McNames, Objective measure of upper extremity motor impairment in Parkinson’s disease with inertial sensors. Conf Proc IEEE Eng Med Biol Soc, 2011. 2011: p. 4378–81.

62. Tsipouras, M.G., et al., An automated methodology for levodopa-induced dyskinesia: assessment based on gyroscope and accelerometer signals. Artif Intell Med, 2012. 55(2): p. 127–35.

63. Griffiths, R.I., et al., Automated assessment of bradykinesia and dyskinesia in Parkinson’s disease. J Parkinsons Dis, 2012. 2(1): p. 47–55.

64. Lee, M.J., et al., Impact of regional striatal dopaminergic function on kinematic parameters of Parkinson’s disease. J Neural Transm (Vienna), 2015. 122(5): p. 669–77.

65. Delrobaei, M., et al., Characterization of multi-joint upper limb movements in a single task to assess bradykinesia. J Neurol Sci, 2016. 368: p. 337–42.

66. Ghassemi, N.H., et al., Combined accelerometer and EMG analysis to differentiate essential tremor from Parkinson’s disease. Conf Proc IEEE Eng Med Biol Soc, 2016. 2016: p. 672–675.

67. Martinez-Manzanera, O., et al., A Method for Automatic and Objective Scoring of Bradykinesia Using Orientation Sensors and Classification Algorithms. IEEE Trans Biomed Eng, 2016. 63(5): p. 1016–1024.

68. Bravo, M., et al. An upper-limbs activities analysis of PD patients in OFF and ON state of medication. in 2016 IEEE Ecuador Technical Chapters Meeting (ETCM). 2016.

69. Tamás, G., et al., Effect of subthalamic stimulation on distal and proximal upper limb movements in Parkinson’s disease. Brain Res, 2016. 1648(Pt A): p. 438–444.

70. Liu, Y., et al., [Quantitative Evaluation of Regularity of Finger Tapping Movement for Patients with Parkinson’s disease]. Sheng Wu Yi Xue Gong Cheng Xue Za Zhi, 2016. 33(5): p. 979–84.

71. Djuric-Jovicic, M., et al., Finger tapping analysis in patients with Parkinson’s disease and atypical parkinsonism. J Clin Neurosci, 2016. 30: p. 49–55.

72. Spasojevic, S., et al., Quantitative Assessment of the Arm/Hand Movements in Parkinson’s Disease Using a Wireless Armband Device. Front Neurol, 2017. 8.

73. Summa, S., et al., Assessing bradykinesia in Parkinson’s disease using gyroscope signals. IEEE Int Conf Rehabil Robot, 2017. 2017: p. 1556–1561.

74. Angeles, P., et al., Automated assessment of symptom severity changes during deep brain stimulation (DBS) therapy for Parkinson’s disease. IEEE Int Conf Rehabil Robot, 2017. 2017: p. 1512–1517.

75. Rabelo, A.G., et al., Objective Assessment of Bradykinesia Estimated from the Wrist Extension in Older Adults and Patients with Parkinson’s Disease. Ann Biomed Eng, 2017. 45(11): p. 2614–2625.

76. Lonini, L., et al., Wearable sensors for Parkinson’s disease: which data are worth collecting for training symptom detection models. npj Digital Medicine, 2018. 1(1): p. 64.

77. Pulliam, C.L., et al., Continuous Assessment of Levodopa Response in Parkinson’s Disease Using Wearable Motion Sensors. IEEE Trans Biomed Eng, 2018. 65(1): p. 159–164.

78. Thomas, I., et al., A Treatment-Response Index From Wearable Sensors for Quantifying Parkinson’s Disease Motor States. IEEE Journal of Biomedical and Health Informatics, 2018. 22(5): p. 1341–1349.

79. Kwon, D.-Y., Y. Kwon, and J.-W. Kim, Quantitative analysis of finger and forearm movements in patients with off state early stage Parkinson’s disease and scans without evidence of dopaminergic deficit (SWEDD). Parkinsonism & Related Disorders, 2018. 57: p. 33–38.

80. di Biase, L., et al., Quantitative Analysis of Bradykinesia and Rigidity in Parkinson’s Disease. Front Neurol, 2018. 9: p. 121.

81. Garza-Rodríguez, A., et al., Pronation and supination analysis based on biomechanical signals from Parkinson’s disease patients. Artif Intell Med, 2018. 84: p. 7–22.

82. Garza-Rodríguez, A., et al., Fuzzy inference model based on triaxial signals for pronation and supination assessment in Parkinson’s disease patients. Artificial Intelligence in Medicine, 2020. 105: p. 101873.

83. Cavallo, F., et al., Upper limb motor pre-clinical assessment in Parkinson’s disease using machine learning. Parkinsonism Relat Disord, 2019. 63: p. 111–116.

84. Bermeo, J., et al., Artificial Neural Network Applied like Qualifier of Symptoms in Patients with Parkinson’s Disease by Evaluating the Movement of Upper-Limbs Activities. 2019. p. 409–414.

85. Li, J., et al., Three-Dimensional Pattern Features in Finger Tapping Test for Patients with Parkinson’s disease. Annu Int Conf IEEE Eng Med Biol Soc, 2020. 2020: p. 3676–3679.

86. Shawen, N., et al., Role of data measurement characteristics in the accurate detection of Parkinson’s disease symptoms using wearable sensors. Journal of NeuroEngineering and Rehabilitation, 2020. 17(1): p. 52.

87. Hssayeni, M.D., et al., Dyskinesia estimation during activities of daily living using wearable motion sensors and deep recurrent networks. Scientific Reports, 2021. 11(1): p. 7865.

88. Park, D.J., et al., Evaluation for Parkinsonian Bradykinesia by deep learning modeling of kinematic parameters. J Neural Transm (Vienna), 2021. 128(2): p. 181–189.

89. Habets, J.G.V., et al., Rapid Dynamic Naturalistic Monitoring of Bradykinesia in Parkinson’s Disease Using a Wrist-Worn Accelerometer. Sensors (Basel), 2021. 21(23).

90. Romano, P., et al., Sensor Network for Analyzing Upper Body Strategies in Parkinson’s Disease versus Normative Kinematic Patterns. Sensors (Basel), 2021. 21(11).

91. Bravo, M., et al. A system for finger tremor quantification in patients with Parkinson’s disease. in 2017 39th Annual International Conference of the IEEE Engineering in Medicine and Biology Society (EMBC). 2017.

92. Chan, P.Y., et al., Motion characteristics of subclinical tremors in Parkinson’s disease and normal subjects. Scientific Reports, 2022. 12(1): p. 4021.

93. Delrobaei, M., et al., Towards remote monitoring of Parkinson’s disease tremor using wearable motion capture systems. J Neurol Sci, 2018. 384: p. 38–45.

94. Makabe, H. and K. Sakamoto, Judgment of disability stages in Parkinson disease patients due to pathological tremor of index finger. Electromyogr Clin Neurophysiol, 2000. 40(7): p. 397–409.

95. Rahimi, F., et al., Effective Management of Upper Limb Parkinsonian Tremor by IncobotulinumtoxinA Injections Using Sensor-based Biomechanical Patterns. Tremor Other Hyperkinet Mov (N Y), 2015. 5: p. 348.

96. Schaefer, L.V., et al., Mechanomyography and acceleration show interlimb asymmetries in Parkinson patients without tremor compared to controls during a unilateral motor task. Scientific Reports, 2021. 11(1): p. 2631.

97. Thanawattano, C., et al., Temporal fluctuation analysis of tremor signal in Parkinson’s disease and Essential tremor subjects. Annu Int Conf IEEE Eng Med Biol Soc, 2015. 2015: p. 6054–7.

98. Lukšys, D., G. Jonaitis, and J. Griškevičius, Quantitative Analysis of Parkinsonian Tremor in a Clinical Setting Using Inertial Measurement Units. Parkinson&#x2019;s Disease, 2018. 2018: p. 1683831.

99. Zhu, N. and N.S. Miller, Assessment System for Parkinson’s Disease Tremor and Correlation Analysis With Applied Signal Processing Algorithms. Journal of Engineering and Science in Medical Diagnostics and Therapy, 2020. 3(4).

100. Rigas, G., et al., Tremor UPDRS estimation in home environment. Annu Int Conf IEEE Eng Med Biol Soc, 2016. 2016: p. 3642–3645.

101. Heldman, D.A., et al., The modified bradykinesia rating scale for Parkinson’s disease: reliability and comparison with kinematic measures. Mov Disord, 2011. 26(10): p. 1859–63.

102. Ali, S.M., et al., Wearable sensors during drawing tasks to measure the severity of essential tremor. Scientific Reports, 2022. 12(1): p. 5242.

103. Benito-León, J., et al., Essential tremor severity and anatomical changes in brain areas controlling movement sequencing. Ann Clin Transl Neurol, 2019. 6(1): p. 83–97.

104. Budini, F., et al., Dexterity training improves manual precision in patients affected by essential tremor. Arch Phys Med Rehabil, 2014. 95(4): p. 705–10.

105. Heo, J.H., et al., Sensory electrical stimulation for suppression of postural tremor in patients with essential tremor. Biomed Mater Eng, 2015. 26 Suppl 1: p. S803–9.

106. McGurrin, P., et al., Quantifying Tremor in Essential Tremor Using Inertial Sensors-Validation of an Algorithm. IEEE J Transl Eng Health Med, 2021. 9: p. 2700110.

107. Chan, P.Y., et al., An In–Laboratory Validity and Reliability Tested System for Quantifying Hand–Arm Tremor in Motions. IEEE Transactions on Neural Systems and Rehabilitation Engineering, 2018. 26(2): p. 460–467.

108. López-Blanco, R., et al., Essential tremor quantification based on the combined use of a smartphone and a smartwatch: The NetMD study. Journal of Neuroscience Methods, 2018. 303: p. 95–102.

109. Šprdlík, O., et al., Tremor analysis by decomposition of acceleration into gravity and inertial acceleration using inertial measurement unit. Biomedical Signal Processing and Control, 2011. 6(3): p. 269–279.

110. Kwon, D.Y., et al., Quantitative measures of postural tremor at the upper limb joints in patients with essential tremor. Technol Health Care, 2020. 28(S1): p. 499–507.

111. Samotus, O., et al., Functional Ability Improved in Essential Tremor by IncobotulinumtoxinA Injections Using Kinematically Determined Biomechanical Patterns – A New Future. PLOS ONE, 2016. 11(4): p. e0153739.

112. Gallego, J., et al., A Multimodal Human-Robot Interface to Drive a Neuroprosthesis for Tremor Management. Systems, Man, and Cybernetics, Part C: Applications and Reviews, IEEE Transactions on, 2012. 42: p. 1159–1168.

113. Heldman, D.A., et al., Essential tremor quantification during activities of daily living. Parkinsonism & related disorders, 2011. 17(7): p. 537–542.

114. Kamper, D.G., et al., Alterations in reaching after stroke and their relation to movement direction and impairment severity. Arch Phys Med Rehabil, 2002. 83(5): p. 702–7.

115. Hester, T., et al. Using wearable sensors to measure motor abilities following stroke. in International Workshop on Wearable and Implantable Body Sensor Networks (BSN’06). 2006.

116. Knorr, B., et al. Quantitative Measures of Functional Upper Limb Movement in Persons after Stroke. in Conference Proceedings. 2nd International IEEE EMBS Conference on Neural Engineering, 2005. 2005.

117. Thies, S.B., et al., Movement variability in stroke patients and controls performing two upper limb functional tasks: a new assessment methodology. J Neuroeng Rehabil, 2009. 6: p. 2.

118. Parnandi, A., E. Wade, and M. Mataric, Motor function assessment using wearable inertial sensors. Conf Proc IEEE Eng Med Biol Soc, 2010. 2010: p. 86–9.

119. Del Din, S., et al., Estimating Fugl-Meyer clinical scores in stroke survivors using wearable sensors. Annu Int Conf IEEE Eng Med Biol Soc, 2011. 2011: p. 5839–42.

120. Patel, S., et al., Tracking motor recovery in stroke survivors undergoing rehabilitation using wearable technology. Annu Int Conf IEEE Eng Med Biol Soc, 2010. 2010: p. 6858–61.

121. van Meulen, F.B., et al., Assessment of daily-life reaching performance after stroke. Ann Biomed Eng, 2015. 43(2): p. 478–86.

122. Otten, P., J. Kim, and S.H. Son, A Framework to Automate Assessment of Upper-Limb Motor Function Impairment: A Feasibility Study. Sensors, 2015. 15(8): p. 20097–20114.

123. Carpinella, I., D. Cattaneo, and M. Ferrarin, Quantitative assessment of upper limb motor function in Multiple Sclerosis using an instrumented Action Research Arm Test. J Neuroeng Rehabil, 2014. 11: p. 67.

124. Carpinella, I., D. Cattaneo, and M. Ferrarin, Hilbert-Huang transform based instrumental assessment of intention tremor in multiple sclerosis. J Neural Eng, 2015. 12(4): p. 046011.

125. Ketteringham, L.P., et al. Measuring Intention Tremor in Multiple Sclerosis using Inertial Measurement Unit (IMU) Devices. in BIODEVICES. 2011.

126. Teufl, S., et al., Quantifying upper limb tremor in people with multiple sclerosis using Fast Fourier Transform based analysis of wrist accelerometer signals. J Rehabil Assist Technol Eng, 2021. 8: p. 2055668320966955.

127. Teufl, S., et al., Objective identification of upper limb tremor in multiple sclerosis using a wrist-worn motion sensor: Establishing validity and reliability. British Journal of Occupational Therapy, 2017. 80(10): p. 596–602.

128. Western, D.G., et al., Personalised profiling to identify clinically relevant changes in tremor due to multiple sclerosis. BMC Medical Informatics and Decision Making, 2019. 19(1): p. 162.

129. Krishna, R., et al., Quantitative assessment of cerebellar ataxia, through automated limb functional tests. Journal of NeuroEngineering and Rehabilitation, 2019. 16(1): p. 31.

130. Uswatte, G., et al., Objective measurement of functional upper-extremity movement using accelerometer recordings transformed with a threshold filter. Stroke, 2000. 31(3): p. 662–7.

131. Nguyen, K.D., et al., Entropy-based analysis of rhythmic tapping for the quantitative assessment of cerebellar ataxia. Biomedical Signal Processing and Control, 2020. 59: p. 101916.

132. Dominguez-Vega, Z.T., et al., Instrumented classification of patients with early onset ataxia or developmental coordination disorder and healthy control children combining information from three upper limb SARA tests. European Journal of Paediatric Neurology, 2021. 34: p. 74–83.

133. Oubre, B., et al., Decomposition of Reaching Movements Enables Detection and Measurement of Ataxia. Cerebellum, 2021. 20(6): p. 811–822.

134. Gupta, A.S., et al., Real-life Wrist Movement Patterns Capture Motor Impairment in Individuals with Ataxia-Telangiectasia. Cerebellum, 2022: p. 1–11.

135. Martinez-Manzanera, O., et al., Instrumented finger-to-nose test classification in children with ataxia or developmental coordination disorder and controls. Clin Biomech (Bristol, Avon), 2018. 60: p. 51–59.

136. Kashyap, B., et al., Objective Assessment of Cerebellar Ataxia: A Comprehensive and Refined Approach. Scientific Reports, 2020. 10(1): p. 9493.

137. Strohrmann, C., et al., Monitoring motor capacity changes of children during rehabilitation using body-worn sensors. Journal of NeuroEngineering and Rehabilitation, 2013. 10(1): p. 83.

138. Kim, D.H., D.H. An, and W.G. Yoo, Measurement of upper limb movement acceleration and functions in children with cerebral palsy. Technol Health Care, 2018. 26(3): p. 429–435.

139. den Hartog, D., et al., Home-Based Measurements of Dystonia in Cerebral Palsy Using Smartphone-Coupled Inertial Sensor Technology and Machine Learning: A Proof-of-Concept Study. Sensors, 2022. 22(12): p. 4386.

140. Legros, A., et al., Accelerometric measurement of involuntary movements during pallidal deep brain stimulation of patients with generalized dystonia. Brain Res Bull, 2004. 64(4): p. 363–9.

141. Bai, L., et al., Quantitative measurement of upper limb motion pre- and post-treatment with Botulinum Toxin. Measurement, 2021. 168: p. 108304.

142. Rahimi, F., et al., Capturing whole-body mobility of patients with Parkinson disease using inertial motion sensors: expected challenges and rewards. Conf Proc IEEE Eng Med Biol Soc, 2011. 2011: p. 5833–8.

143. Ghassemi, M., et al., Bradykinesia in patients with Parkinson’s disease having levodopa-induced dyskinesias. Brain Res Bull, 2006. 69(5): p. 512–8.

144. Tran, H., et al., A comprehensive scheme for the objective upper body assessments of subjects with cerebellar ataxia. J Neuroeng Rehabil, 2020. 17(1): p. 162.

145. Patel, S., et al., Using wearable sensors to predict the severity of symptoms and motor complications in late stage Parkinson’s Disease. Conf Proc IEEE Eng Med Biol Soc, 2008. 2008: p. 3686–9.

146. Liu, X., et al., Quantifying drug-induced dyskinesias in the arms using digitised spiral-drawing tasks. J Neurosci Methods, 2005. 144(1): p. 47–52.

147. Feigin, V.L., et al., World Stroke Organization (WSO): Global Stroke Fact Sheet 2022. International Journal of Stroke, 2022. 17(1): p. 18–29.

148. Song, P., et al., The global prevalence of essential tremor, with emphasis on age and sex: A meta-analysis. Journal of global health, 2021. 11: p. 04028–04028.

149. McIntyre, S., et al., Cerebral palsy--don’t delay. Dev Disabil Res Rev, 2011. 17(2): p. 114–29.

150. Marras, C., et al., Prevalence of Parkinson’s disease across North America. npj Parkinson’s Disease, 2018. 4(1): p. 21.

151. Okubadejo, N.U., et al., Parkinson’s disease in Africa: A systematic review of epidemiologic and genetic studies. Mov Disord, 2006. 21(12): p. 2150–6.

152. Van Den Eeden, S.K., et al., Incidence of Parkinson’s disease: variation by age, gender, and race/ethnicity. Am J Epidemiol, 2003. 157(11): p. 1015–22.

153. Musselman, K.E., et al., Prevalence of ataxia in children: a systematic review. Neurology, 2014. 82(1): p. 80–89.

154. Ruano, L., et al., The Global Epidemiology of Hereditary Ataxia and Spastic Paraplegia: A Systematic Review of Prevalence Studies. Neuroepidemiology, 2014. 42(3): p. 174–183.

155. Medina, A., et al., Prevalence and Incidence of Huntington’s Disease: An Updated Systematic Review and Meta-Analysis. Movement Disorders. n/a(n/a).

156. Crowell, V., et al., Modeling Manifest Huntington’s Disease Prevalence Using Diagnosed Incidence and Survival Time. Neuroepidemiology, 2021. 55(5): p. 361–368.

157. Del Din, S., et al., Body-Worn Sensors for Remote Monitoring of Parkinson’s Disease Motor Symptoms: Vision, State of the Art, and Challenges Ahead. Journal of Parkinson’s Disease, 2021. 11: p. S35–S47.

158. Jankovic, J., Motor fluctuations and dyskinesias in Parkinson’s disease: clinical manifestations. Mov Disord, 2005. 20 Suppl 11: p. S11–6.

159. Serrano, J.I., et al., Identification of activities of daily living in tremorous patients using inertial sensors. Expert Systems with Applications, 2017. 83: p. 40–48.

160. Nguyen, H., et al., Auto detection and segmentation of daily living activities during a Timed Up and Go task in people with Parkinson’s disease using multiple inertial sensors. J Neuroeng Rehabil, 2017. 14(1): p. 26.

161. Pham, M.H., et al., Algorithm for Turning Detection and Analysis Validated under Home-Like Conditions in Patients with Parkinson’s Disease and Older Adults using a 6 Degree-of-Freedom Inertial Measurement Unit at the Lower Back. Front Neurol, 2017. 8: p. 135.

162. Beani, E., et al., Actigraph assessment for measuring upper limb activity in unilateral cerebral palsy. J Neuroeng Rehabil, 2019. 16(1): p. 30.

163. Ahmadi, M.N., et al., Machine Learning to Quantify Physical Activity in Children with Cerebral Palsy: Comparison of Group, Group-Personalized, and Fully-Personalized Activity Classification Models. Sensors (Basel), 2020. 20(14).

164. Braito, I., et al., Assessment of upper limb use in children with typical development and neurodevelopmental disorders by inertial sensors: a systematic review. J Neuroeng Rehabil, 2018. 15(1): p. 94.

165. Biswas, D., et al., Recognizing upper limb movements with wrist worn inertial sensors using k-means clustering classification. Hum Mov Sci, 2015. 40: p. 59–76.

