## Supplementary Material S1 for "Assessment of upper limb movement disorders using wearable sensors during functional tasks: a systematic review"

| **Study ID** | **Reason of exclusion** |
| --- | --- |
| Syeda et al., 2022 [1] | Glove instead of sensors |
| Albani et al., 2019 [2] | Glove instead of sensors |
| Zoccolillo et al., 2015 [3] | Kinect |
| Machado et al., 2016 [4] | Glove instead of sensors |
| Rech et al., 2020 [5] | No wearable sensors |
| Liu et al., 2005 [6] | No wearable sensors |
| Reilmann et al., 2011 [7] | No wearable sensors |
| Nguyen et al., 2020 [8] | No wearable sensors |
| Einarsson et al., 2018 [9] | Kinect |
| Summa et al., 2020 [10] | Kinect |
| Krishna et al., 2021 [11] | No wearable sensors |
| Kishimoto et al., 2022 [12] | No wearable sensors |
| Romano et al., 2022 [13] | No wearable sensors |
| Avanzino et al., 2009 [14] | Glove instead of sensors |
| Blumrosen et al., 2012 [15] | No wearable sensors |
| Duval & Norton, 2006 [16] | No wearable sensors |
| Geiger et al., 2018 [17] | Motion capture system |
| Héroux & Norman, 2011 [18] | No wearable sensors |
| Héroux et al., 2006 [19] | No wearable sensors |
| Khwaounjoo et al., [20] | Camera-based system |
| Meshack & Norman [21] | Laser sensors |
| Milano et al., 2021 [22] | No wearable sensors |
| Mittal et al., 2020 [23] | No wearable sensors |
| Oliveira et al., 2019 [24] | No wearable sensors |
| Papapetropoulos et al., 2008 [25] | No wearable sensors |
| Raethjen et al., 2005 [26] | No wearable sensors |
| Rocon et al., 2010 [27] | No wearable sensors |
| Rozman et al., 2007 [28] | No wearable sensors |
| Samotus et al., 2018 [29] | No acceleration & angular velocity from wearable sensors |
| Samotus et al., 2017 [30] | No acceleration & angular velocity from wearable sensors |
| Schaefer & Bittmann, 2020 [31] | No wearable sensors |
| Lin et al., 2021 [32] | No wearable sensors |
| Zhou et al., 2021 [33] | Glove instead of sensors |
| Daneault et al., 2010 [34] | Laser sensors |
| Kavanagh et al., 2013 [35] | No wearable sensors |
| Grimaldi et al., 2008 [36] | No wearable sensors |
| Dideriksen et al., 2011 [37] | No wearable sensors |
| Kavindya et al., 2020 [38] | Glove instead of sensors |
| Kim et al., 2022 [39] | No wearable sensors |
| Vanitha & Talasila, 2022 [40] | Glove instead of sensors |
| Takanokura & Sakamoto, 2005 [41] | No wearable sensors |
| Alper et al., 2020 [42] | No wearable sensors |
| Amarantini et al., 2022 [43] | No wearable sensors |
| Carignan et al., 2010 [44] | Laser sensors |
| Ferencik et al., 2020 [45] | No wearable sensors |
| Cernera et al., 2021 [46] | Only EMG sensors |
| Alam et al., 2017 [47] | No movement disorder |
| Cahill-Rowley & Rose, 2018 [48] | No movement disorder |
| Enokinozo et al., 2020 [49] | No movement disorder |
| Wade et al., 2010 [50] | No movement disorder |
| Kaneko et al., 2015 [51] | No movement disorder |
| Schasfoort et al., 2006 [52] | No movement disorder |
| Carmona-Almazan et al., 2021 [53] | No movement disorder |
| Jeonghee et al., 2016 [54] | No movement disorder |
| Aswathy et al., 2016 [55] | No movement disorder |
| Aljihmani et al., 2020 [56] | No movement disorder |
| Davidson & Charles, 2017 [57] | No movement disorder |
| Herrnstadt & Menon, 2016 [58] | No movement disorder |
| Angeles et al., 2016 [59] | No movement disorder |
| Teoderescu et al., [60] | No movement disorder |
| Carignan et al., 2012 [61] | No movement disorder |
| Liu et al., 2020 [62] | No movement disorder |
| Lopes et al., 2020 [63] | No movement disorder |
| Gordon et al., 2019 [64] | No specific features |
| Pfister et al., 2020 [65] | No specific features |
| Keinan et al., 2020 [66] | No specific features |
| Roy et al., 2013 [67] | No specific features |
| Campolo et al., 2008 [68] | No specific features |
| Gutowski & Chmielewski [69] | No specific features |
| Velazquez-Pérez et al., 2021 [70] | No specific features |
| Bermeo et al., 2016 [71] | No specific features |
| Bo et al., 2011 [72] | No specific features |
| Lambrecht et al., 2015 [73] | No specific features |
| McNames et al., 2019 [74] | No specific features |
| O’Keefe et al., 2020 [75] | No specific features |
| Tatinati et al., 2013 [76] | No specific features |
| Kalaiarasi & Kumar [77] | No specific features |
| Widjaja et al., 2007 [78] | No specific features |
| Shahtalebi et al., 2020 [79] | No specific features |
| Ibrahim et al., 2021 [80] | No specific features |
| Mahadevan et al., 2020 [81] | No upper limb tasks |
| Evers et al., 2020 [82] | No upper limb tasks |
| Gour et al., 2007 [83] | No upper limb tasks |
| Ilias et al., 2017 [84] | No upper limb tasks |
| Popovic et al., 2011 [85] | No upper limb tasks |
| Liu et al., 2022 [86] | No upper limb tasks |
| Biswas et al., 2014 [87] | Activity recognition |
| Sokal et al., 2015 [88] | Arm use |
| Gordon et al., 2007 [89] | Arm use |
| Giuffrida et al., 2008 [90] | Activity recognition |
| Shah et al., 2017 [91] | Passive movements |
| Serrano et al., 2017 [92] | Activity recognition |
| Wang et al., 2019 [93] | Activity recognition |
| Datta et al., 2020 [94] | Arm use |
| Brogioli et al., 2017 [95] | Activity recognition |
| Munoz-Organero et al., 2019 [96] | Activity recognition |
| Munoz-Organero et al., 2018 [97] | Activity recognition |
| Poitras et al., 2022 [98] | Arm use |
| Merlo et al., 2021 [99] | Arm use |
| Ponsiglione et al., 2022 [100] | Sensor location not specified |
| Khodakarami et al., 2019 [101] | Same features as previously included study and author |
| Aghanavesi et al., 2020 [102] | Same features as previously included study and author |
| Heldman et al., 2017 [103] | Same features as previously included study and author |
| Bobic et al., 2019 [104] | Same features as previously included study and author |

References

1.	Syeda, H.B., et al., *Amplitude setting and dopamine response of finger tapping and gait are related in Parkinson’s disease.* Scientific Reports, 2022. **12**(1): p. 4180.

2.	Albani, G., et al., *An Integrated Multi-Sensor Approach for the Remote Monitoring of Parkinson’s Disease.* Sensors, 2019. **19**(21): p. 4764.

3.	Zoccolillo, L., et al., *Video-game based therapy performed by children with cerebral palsy: a cross-over randomized controlled trial and a cross-sectional quantitative measure of physical activity.* Eur J Phys Rehabil Med, 2015. **51**(6): p. 669-76.

4.	Machado, A.R., et al., *Feature visualization and classification for the discrimination between individuals with Parkinson's disease under levodopa and DBS treatments.* Biomed Eng Online, 2016. **15**(1): p. 169.

5.	Rech, K.D., et al., *Fugl-Meyer Assessment Scores Are Related With Kinematic Measures in People with Chronic Hemiparesis after Stroke.* J Stroke Cerebrovasc Dis, 2020. **29**(1): p. 104463.

6.	Liu, X., et al., *Quantifying drug-induced dyskinesias in the arms using digitised spiral-drawing tasks.* J Neurosci Methods, 2005. **144**(1): p. 47-52.

7.	Reilmann, R., et al., *Assessment of involuntary choreatic movements in Huntington's disease--toward objective and quantitative measures.* Mov Disord, 2011. **26**(12): p. 2267-73.

8.	Nguyen, K.D., et al., *The Assessment of Upper Limb Functionality in Friedreich Ataxia via Self-Feeding Activity.* IEEE Trans Neural Syst Rehabil Eng, 2020. **28**(4): p. 924-933.

9.	Einarsson, G., et al. *Computer Aided Identification of Motion Disturbances Related to Parkinson’s Disease*. in *PRedictive Intelligence in MEdicine*. 2018. Cham: Springer International Publishing.

10.	Summa, S., et al., *Development of SaraHome: A novel, well-accepted, technology-based assessment tool for patients with ataxia.* Computer Methods and Programs in Biomedicine, 2020. **188**: p. 105257.

11.	Krishna, R., et al., *Quantitative Assessment of Friedreich Ataxia via Self-Drinking Activity.* IEEE J Biomed Health Inform, 2021. **25**(6): p. 1985-1996.

12.	Kishimoto, Y., et al., *Quantitative evaluation of upper limb ataxia in spinocerebellar ataxias.* Ann Clin Transl Neurol, 2022. **9**(4): p. 529-539.

13.	Romano, A., et al., *Upper Body Physical Rehabilitation for Children with Ataxia through IMU-Based Exergame.* J Clin Med, 2022. **11**(4).

14.	Avanzino, L., et al., *Cerebellar involvement in timing accuracy of rhythmic finger movements in essential tremor.* Eur J Neurosci, 2009. **30**(10): p. 1971-9.

15.	Blumrosen, G., et al., *Noncontact tremor characterization using low-power wideband radar technology.* IEEE Trans Biomed Eng, 2012. **59**(3): p. 674-86.

16.	Duval, C. and L. Norton, *Tremor in patients with migraine.* Headache, 2006. **46**(6): p. 1005-10.

17.	Geiger, D.W., D.L. Eggett, and S.K. Charles, *A method for characterizing essential tremor from the shoulder to the wrist.* Clinical Biomechanics, 2018. **52**: p. 117-123.

18.	Héroux, M.E., G. Pari, and K.E. Norman, *The effect of inertial loading on wrist kinetic tremor and rhythmic muscle activity in individuals with essential tremor.* Clin Neurophysiol, 2011. **122**(9): p. 1794-801.

19.	Héroux, M.E., et al., *Upper-extremity disability in essential tremor.* Arch Phys Med Rehabil, 2006. **87**(5): p. 661-70.

20.	Khwaounjoo, P., et al., *Non-Contact Hand Movement Analysis for Optimal Configuration of Smart Sensors to Capture Parkinson's Disease Hand Tremor.* Sensors (Basel), 2022. **22**(12).

21.	Meshack, R.P. and K.E. Norman, *A randomized controlled trial of the effects of weights on amplitude and frequency of postural hand tremor in people with Parkinson's disease.* Clin Rehabil, 2002. **16**(5): p. 481-92.

22.	Milano, F., et al., *Parkinson’s Disease Patient Monitoring: A Real-Time Tracking and Tremor Detection System Based on Magnetic Measurements.* Sensors, 2021. **21**(12): p. 4196.

23.	Mittal, S.O., et al., *Novel Botulinum Toxin Injection Protocols for Parkinson Tremor and Essential Tremor - the Yale Technique and Sensor-Based Kinematics Procedure for Safe and Effective Treatment.* Tremor Other Hyperkinet Mov (N Y), 2020. **10**: p. 61.

24.	Oliveira, F.H.M., et al., *On the Use of Non-Contact Capacitive Sensors for the Assessment of Postural Hand Tremor of Individuals with Parkinson's Disease.* Annu Int Conf IEEE Eng Med Biol Soc, 2019. **2019**: p. 6591-6594.

25.	Papapetropoulos, S., et al., *Objective monitoring of tremor and bradykinesia during DBS surgery for Parkinson disease.* Neurology, 2008. **70**(15): p. 1244-9.

26.	Raethjen, J., et al., *Parkinsonian action tremor: interference with object manipulation and lacking levodopa response.* Exp Neurol, 2005. **194**(1): p. 151-60.

27.	Rocon, E., et al., *Multimodal BCI-mediated FES suppression of pathological tremor.* Annu Int Conf IEEE Eng Med Biol Soc, 2010. **2010**: p. 3337-40.

28.	Rozman, J., A. Bartolić, and S. Ribaric, *A new method for selective measurement of joint movement in hand tremor in Parkinson's disease patients.* J Med Eng Technol, 2007. **31**(4): p. 305-11.

29.	Samotus, O., et al., *Botulinum Toxin Type A Injections as Monotherapy for Upper Limb Essential Tremor Using Kinematics.* Can J Neurol Sci, 2018. **45**(1): p. 11-22.

30.	Samotus, O., J. Lee, and M. Jog, *Long-term tremor therapy for Parkinson and essential tremor with sensor-guided botulinum toxin type A injections.* PLoS One, 2017. **12**(6): p. e0178670.

31.	Schaefer, L.V. and F.N. Bittmann, *Parkinson patients without tremor show changed patterns of mechanical muscle oscillations during a specific bilateral motor task compared to controls.* Scientific reports, 2020. **10**(1): p. 1168-1168.

32.	Lin, C.H., et al., *Tremor Class Scaling for Parkinson Disease Patients Using an Array X-Band Microwave Doppler-Based Upper Limb Movement Quantizer.* IEEE Sensors Journal, 2021. **21**(19): p. 21473-21485.

33.	Zhou, Y., et al., *Analysis of the Effect of Common Disturbances on the Safety of a Wearable Tremor Suppression Device.* IEEE Robotics and Automation Letters, 2021. **6**(2): p. 2846-2853.

34.	Daneault, J.F., B. Carignan, and C. Duval, *Bilateral effect of a unilateral voluntary modulation of physiological tremor.* Clin Neurophysiol, 2010. **121**(5): p. 734-43.

35.	Kavanagh, J.J., et al., *Bilateral tremor responses to unilateral loading and fatiguing muscle contractions.* J Neurophysiol, 2013. **110**(2): p. 431-40.

36.	Grimaldi, G., et al., *Effects of inertia and wrist oscillations on contralateral neurological postural tremor using the wristalyzer, a new myohaptic device.* IEEE Trans Biomed Circuits Syst, 2008. **2**(4): p. 269-79.

37.	Dideriksen, J.L., et al., *EMG-based characterization of pathological tremor using the iterated Hilbert transform.* IEEE Trans Biomed Eng, 2011. **58**(10): p. 2911-21.

38.	Kavindya, P., et al. *Evaluation of Hand Tremor Frequency Among Patients in Sri Lanka using a Soft Glove*. in *2020 Moratuwa Engineering Research Conference (MERCon)*. 2020.

39.	Kim, J., et al., *Fitts' Law Based Performance Metrics to Quantify Tremor in Individuals With Essential Tremor.* IEEE J Biomed Health Inform, 2022. **26**(5): p. 2169-2179.

40.	Vanitha, K. and V. Talasila, *Machine Learning Techniques for Automated Tremor Detection in the Presence of External Stressors.* International Journal of Circuits, Systems and Signal Processing, 2022. **16**: p. 551-560.

41.	Takanokura, M. and K. Sakamoto, *Neuromuscular control of physiological tremor during elastic load.* Med Sci Monit, 2005. **11**(4): p. Cr143-52.

42.	Alper, M.A., J. Goudreau, and M. Daniel, *Pose and Optical Flow Fusion (POFF) for accurate tremor detection and quantification.* Biocybernetics and Biomedical Engineering, 2020. **40**(1): p. 468-481.

43.	Amarantini, D., et al., *Quantification of Head Tremors in Medical Conditions: A Comparison of Analyses Using a 2D Video Camera and a 3D Wireless Inertial Motion Unit.* Sensors (Basel), 2022. **22**(6).

44.	Carignan, B., J.F. Daneault, and C. Duval, *Quantifying the importance of high frequency components on the amplitude of physiological tremor.* Exp Brain Res, 2010. **202**(2): p. 299-306.

45.	Ferenčík, N., et al., *The Rehapiano-Detecting, Measuring, and Analyzing Action Tremor Using Strain Gauges.* Sensors (Basel), 2020. **20**(3).

46.	Cernera, S., et al., *Wearable sensor-driven responsive deep brain stimulation for essential tremor.* Brain Stimul, 2021. **14**(6): p. 1434-1443.

47.	Alam, M.N., et al. *A Quantitative Assessment of Bradykinesia Using Inertial Measurement Unit*. in *2017 Design of Medical Devices Conference*. 2017.

48.	Cahill-Rowley, K. and J. Rose, *Temporal-spatial reach parameters derived from inertial sensors correlate to neurodevelopment in toddlers born preterm.* J Biomech, 2018. **72**: p. 17-22.

49.	Enokizono, T., et al., *Quantitative assessment of fine motor skills in children using magnetic sensors.* Brain and Development, 2020. **42**(6): p. 421-430.

50.	Wade, E., A.R. Parnandi, and M.J. Mataríc. *Automated administration of the Wolf Motor Function Test for post-stroke assessment*. in *2010 4th International Conference on Pervasive Computing Technologies for Healthcare*. 2010.

51.	Kaneko, M., et al., *Soft Neurological Signs in Childhood by Measurement of Arm Movements Using Acceleration and Angular Velocity Sensors.* Sensors, 2015. **15**(10): p. 25793-25808.

52.	Schasfoort, F.C., et al., *Objective measurement of upper limb activity and mobility during everyday behavior using ambulatory accelerometry: the upper limb activity monitor.* Behav Res Methods, 2006. **38**(3): p. 439-46.

53.	Carmona-Almazan, A., et al., *Triaxial Accelerometry Wireless System for Characterization of Parkinsonian Tremor.* Annu Int Conf IEEE Eng Med Biol Soc, 2021. **2021**: p. 7320-7323.

54.	Jeonghee, K., et al., *Longitudinal wearable tremor measurement system with activity recognition algorithms for upper limb tremor.* Annu Int Conf IEEE Eng Med Biol Soc, 2016. **2016**: p. 6166-6169.

55.	Aswathy, K.P., A. Kalaiarasi, and L.A. Kumar. *Central mechanism of tremor suppression at musculoskeletal level*. in *2016 International Conference on Electrical, Electronics, and Optimization Techniques (ICEEOT)*. 2016.

56.	Aljihmani, L., et al. *Detection of Tremor Associated with Rest and Effort Activity Using Machine Learning*. in *2020 International Conference Automatics and Informatics (ICAI)*. 2020.

57.	Davidson, A.D. and S.K. Charles, *Fundamental Principles of Tremor Propagation in the Upper Limb.* Annals of biomedical engineering, 2017. **45**(4): p. 1133-1147.

58.	Herrnstadt, G. and C. Menon, *Voluntary-Driven Elbow Orthosis with Speed-Controlled Tremor Suppression.* Frontiers in bioengineering and biotechnology, 2016. **4**: p. 29-29.

59.	Angeles, P., et al. *A Wearable Automated System to Quantify Parkinsonian Symptoms Enabling Closed Loop Deep Brain Stimulation*. in *Towards Autonomous Robotic Systems*. 2016. Cham: Springer International Publishing.

60.	Teodorescu, H.-N.L., et al., *Fuzzy methods in tremor assessment, prediction, and rehabilitation.* Artificial Intelligence in Medicine, 2001. **21**(1): p. 107-130.

61.	Carignan, B., J.F. Daneault, and C. Duval, *The organization of upper limb physiological tremor.* Eur J Appl Physiol, 2012. **112**(4): p. 1269-84.

62.	Liu, W., T. Kai, and K. Kiguchi, *Tremor Suppression With Mechanical Vibration Stimulation.* IEEE Access, 2020. **8**: p. 226199-226212.

63.	Lopes, E.M., et al., *iHandU: A Novel Quantitative Wrist Rigidity Evaluation Device for Deep Brain Stimulation Surgery.* Sensors (Basel, Switzerland), 2020. **20**(2): p. 331.

64.	Gordon, Mark F., et al., *Quantification of Motor Function in Huntington Disease Patients Using Wearable Sensor Devices.* Digital Biomarkers, 2019. **3**: p. 103-115.

65.	Pfister, F.M.J., et al., *High-Resolution Motor State Detection in Parkinson’s Disease Using Convolutional Neural Networks.* Scientific Reports, 2020. **10**(1): p. 5860.

66.	Keinan, A., T. Bar-Shalita, and S. Portnoy, *An Instrumented Assessment of a Rhythmic Finger Task among Children with Motor Coordination Difficulties.* Sensors, 2020. **20**(16): p. 4554.

67.	Roy, S.H., et al., *High-resolution tracking of motor disorders in Parkinson's disease during unconstrained activity.* Mov Disord, 2013. **28**(8): p. 1080-7.

68.	Campolo, D., et al., *A novel technological approach towards the early diagnosis of neurodevelopmental disorders.* Annu Int Conf IEEE Eng Med Biol Soc, 2008. **2008**: p. 4875-8.

69.	Gutowski, T. and M. Chmielewski, *An Algorithmic Approach for Quantitative Evaluation of Parkinson’s Disease Symptoms and Medical Treatment Utilizing Wearables and Multi-Criteria Symptoms Assessment.* IEEE Access, 2021. **9**: p. 24133-24144.

70.	Velázquez-Pérez, L., et al., *Prodromal Spinocerebellar Ataxia Type 2 Subjects Have Quantifiable Gait and Postural Sway Deficits.* Mov Disord, 2021. **36**(2): p. 471-480.

71.	Bravo, M., et al. *A system for finger tremor quantification in patients with Parkinson's disease*. in *2017 39th Annual International Conference of the IEEE Engineering in Medicine and Biology Society (EMBC)*. 2017.

72.	A. P. L, B., P. Poignet, and C. Geny, *Pathological Tremor and Voluntary Motion Modeling and Online Estimation for Active Compensation.* IEEE Transactions on Neural Systems and Rehabilitation Engineering, 2011. **19**(2): p. 177-185.

73.	Lambrecht, S., et al., *Automatic real-time monitoring and assessment of tremor parameters in the upper limb from orientation data.* Frontiers in Neuroscience, 2014. **8**.

74.	McNames, J., et al. *A Two-Stage Tremor Detection Algorithm for Wearable Inertial Sensors During Normal Daily Activities*. in *2019 41st Annual International Conference of the IEEE Engineering in Medicine and Biology Society (EMBC)*. 2019.

75.	O'Keefe, J.A., et al., *Prodromal Markers of Upper Limb Deficits in FMR1 Premutation Carriers and Quantitative Outcome Measures for Future Clinical Trials in Fragile X-associated Tremor/Ataxia Syndrome.* Mov Disord Clin Pract, 2020. **7**(7): p. 810-819.

76.	Tatinati, S., et al. *Online LS-SVM based multi-step prediction of physiological tremor for surgical robotics*. in *2013 35th Annual International Conference of the IEEE Engineering in Medicine and Biology Society (EMBC)*. 2013.

77.	Kalaiarasi, A. and L.A. Kumar, *Sensor Based Portable Tremor Suppression Device for Stroke Patients.* Acupuncture & Electro-Therapeutics Research, 2018. **43**(1): p. 29-37.

78.	Widjaja, F., et al. *Towards a sensing system for quantification of pathological tremor*. in *2007 International Conference on Intelligent and Advanced Systems*. 2007.

79.	Shahtalebi, S., et al., *PHTNet: Characterization and Deep Mining of Involuntary Pathological Hand Tremor using Recurrent Neural Network Models.* Sci Rep, 2020. **10**(1): p. 2195.

80.	Ibrahim, A., et al., *Real-Time Voluntary Motion Prediction and Parkinson's Tremor Reduction Using Deep Neural Networks.* IEEE Trans Neural Syst Rehabil Eng, 2021. **29**: p. 1413-1423.

81.	Mahadevan, N., et al., *Development of digital biomarkers for resting tremor and bradykinesia using a wrist-worn wearable device.* npj Digital Medicine, 2020. **3**(1): p. 5.

82.	Evers, L.J., et al., *Real-Life Gait Performance as a Digital Biomarker for Motor Fluctuations: The Parkinson@Home Validation Study.* J Med Internet Res, 2020. **22**(10): p. e19068.

83.	Gour, J., et al., *Movement patterns of peak-dose levodopa-induced dyskinesias in patients with Parkinson's disease.* Brain Res Bull, 2007. **74**(1-3): p. 66-74.

84.	Ilias, T., et al. *Using measurements from wearable sensors for automatic scoring of Parkinson's disease motor states: Results from 7 patients*. in *2017 39th Annual International Conference of the IEEE Engineering in Medicine and Biology Society (EMBC)*. 2017.

85.	Popović Maneski, L., et al., *Electrical stimulation for the suppression of pathological tremor.* Med Biol Eng Comput, 2011. **49**(10): p. 1187-93.

86.	Liu, S., et al., *Comprehensive analysis of resting tremor based on acceleration signals of patients with Parkinson's disease.* Technol Health Care, 2022. **30**(4): p. 895-907.

87.	Biswas, D., et al., *Recognizing upper limb movements with wrist worn inertial sensors using k-means clustering classification.* Hum Mov Sci, 2015. **40**: p. 59-76.

88.	Sokal, B., et al., *Everyday movement and use of the arms: Relationship in children with hemiparesis differs from adults.* J Pediatr Rehabil Med, 2015. **8**(3): p. 197-206.

89.	Gordon, A.M., et al., *Efficacy of a hand-arm bimanual intensive therapy (HABIT) in children with hemiplegic cerebral palsy: a randomized control trial.* Dev Med Child Neurol, 2007. **49**(11): p. 830-8.

90.	Giuffrida, J.P., et al., *Upper-extremity stroke therapy task discrimination using motion sensors and electromyography.* IEEE Trans Neural Syst Rehabil Eng, 2008. **16**(1): p. 82-90.

91.	Shah, A., et al., *A novel assistive method for rigidity evaluation during deep brain stimulation surgery using acceleration sensors.* J Neurosurg, 2017. **127**(3): p. 602-612.

92.	Serrano, J.I., et al., *Identification of activities of daily living in tremorous patients using inertial sensors.* Expert Systems with Applications, 2017. **83**: p. 40-48.

93.	Wang, H., M. Irfan, and B.-J. Beijnum, *Measuring Upper-Extremity Use with One IMU*. 2019. 93-100.

94.	Datta, S., et al., *Automated Scoring of Hemiparesis in Acute Stroke From Measures of Upper Limb Co-Ordination Using Wearable Accelerometry.* IEEE Trans Neural Syst Rehabil Eng, 2020. **28**(4): p. 805-816.

95.	Brogioli, M., et al., *Multi-Day Recordings of Wearable Sensors Are Valid and Sensitive Measures of Function and Independence in Human Spinal Cord Injury.* J Neurotrauma, 2017. **34**(6): p. 1141-1148.

96.	Muñoz-Organero, M., et al., *Using Recurrent Neural Networks to Compare Movement Patterns in ADHD and Normally Developing Children Based on Acceleration Signals from the Wrist and Ankle.* Sensors (Basel), 2019. **19**(13).

97.	Muñoz-Organero, M., et al., *Automatic Extraction and Detection of Characteristic Movement Patterns in Children with ADHD Based on a Convolutional Neural Network (CNN) and Acceleration Images.* Sensors (Basel), 2018. **18**(11).

98.	Poitras, I., et al., *Accelerometry-Based Metrics to Evaluate the Relative Use of the More Affected Arm during Daily Activities in Adults Living with Cerebral Palsy.* Sensors, 2022. **22**(3): p. 1022.

99.	Merlo, A., et al., *Monitoring Involuntary Muscle Activity in Acute Patients with Upper Motor Neuron Lesion by Wearable Sensors: A Feasibility Study.* Sensors, 2021. **21**(9): p. 3120.

100.	Ponsiglione, A.M., et al., *Statistical Analysis and Kinematic Assessment of Upper Limb Reaching Task in Parkinson's Disease.* Sensors (Basel), 2022. **22**(5).

101.	Khodakarami, H., et al., *Prediction of the Levodopa Challenge Test in Parkinson's Disease Using Data from a Wrist-Worn Sensor.* Sensors (Basel), 2019. **19**(23).

102.	Aghanavesi, S., et al., *A multiple motion sensors index for motor state quantification in Parkinson's disease.* Comput Methods Programs Biomed, 2020. **189**: p. 105309.

103.	Heldman, D.A., et al., *App-Based Bradykinesia Tasks for Clinic and Home Assessment in Parkinson's Disease: Reliability and Responsiveness.* J Parkinsons Dis, 2017. **7**(4): p. 741-747.

104.	Bobić, V., et al., *An Expert System for Quantification of Bradykinesia Based on Wearable Inertial Sensors.* Sensors (Basel), 2019. **19**(11).
